# Circulating Cell-Free DNA Methylation Profiles Enable Disease-Specific Detection of Alzheimer’s, Parkinson’s, and ALS from Blood

**DOI:** 10.1101/2025.10.07.25337503

**Authors:** Chad Pollard, Erin Saito, Ryan Miller, Isaac Sirland, Mykle Keni, Andrew Jenkins, Jonathon T. Hill, Tim Jenkins

**Author notes:** Correspondence: Jonathon Hill, Department of Cell Biology & Physiology, 4005 LSB, Brigham Young University, Provo, UT 84602, USA.

## Abstract

Noninvasive biomarkers for neurodegenerative diseases are urgently needed for earlier detection, monitoring, and intervention. To address this, we developed a blood-based cfDNA methylation platform. A whole-genome nanopore methylation atlas of six primary human neural cell types—cortical, dopaminergic, and spinal motor neurons, astrocytes, Schwann cells, microglia—was used to train classifiers that assign cfDNA to its neuronal or glial origin. Classifiers were validated *in silico* and applied to 219 plasma samples from patients with Alzheimer’s disease (AD), Parkinson’s disease (PD), amyotrophic lateral sclerosis (ALS), and controls. Cortical cfDNA was elevated in AD, dopaminergic cfDNA in PD, and spinal motor neuron cfDNA in ALS, with predictive modeling achieving AUCs >0.98. In mild cognitive impairment, cortical cfDNA elevations identified individuals who later progressed to AD, supporting predictive utility. A multivariate model integrating multiple neurons improved accuracy, suggesting cfDNA methylation profiling as a scalable framework for noninvasive neurodegeneration detection and monitoring.

## 1. INTRODUCTION

Neurodegenerative diseases constitute one of the fastest-growing public health challenges worldwide. Alzheimer’s disease (AD) alone currently affects ~7 million individuals over age 65 in the U.S., with prevalence projected to double by 2060^1^. Parkinson’s disease (PD) impacts millions globally^2,3^, and approximately 30,000 Americans live with amyotrophic lateral sclerosis (ALS)^4,5^. These diseases cause progressive and irreversible loss of neurons, leading to cognitive impairment, motor dysfunction, and premature mortality.

Despite decades of research, current diagnostics remain fundamentally reactive, applied in advanced disease stages after substantial neuron loss^6–8^, when therapeutic interventions are limited and prognosis is poor^9^. Clinical symptoms, cerebrospinal fluid (CSF) analysis, and neuroimaging (MRI, PET) can confirm advanced disease but are invasive, expensive, and inaccessible for early screening or routine, longitudinal use^10–12^.

Blood-based protein biomarkers such as NfL, GFAP, amyloid-β, tau, and α-synuclein are improving accessibility but remain insufficient for early screening or disease monitoring. General markers of neurodegeneration signal neuronal injury without resolving which cells or brain regions are affected^13,14^. Diagnostic protein biomarkers (e.g., amyloid in AD, α-synuclein in PD) are narrow in scope, limited in their ability to generalize across conditions, and often fail to correlate with disease progression^13,15^. Recent updates to AD diagnostic criteria underscore this limitation. While amyloid and tau biomarkers can confirm AD neuropathology, they are not recommended for presymptomatic use due to limited sensitivity and the current lack of effective presymptomatic disease-modifying therapies^15^.

The result is a diagnostic catch-22: biomarkers confirm disease, but only in advanced disease stages. Development of early detection tools has been sidelined because they cannot yet demonstrate meaningful clinical impact. As a result, clinicians still lack scalable tools to detect and track active neurodegeneration in real time, when intervention would matter most.

Selective neuronal cell death is a fundamental hallmark of neurodegenerative diseases^16^, yet current protein biomarkers provide only indirect or nonspecific readouts of this pathology. Circulating cell-free DNA (cfDNA) offers a fundamentally different class of biomarkers that may address these challenges. Fragments of DNA released into blood during neuronal apoptosis carry stable^17,18^, “cell-of-origin” methylation signatures of the dying cell. Thus cfDNA presents a biologically anchored, real-time measure of active cell loss due to cfDNA’s short circulating half-life^19^. Importantly, this approach aims to monitor a defining feature of neurodegenerative disease—the loss of neurons—providing unique insights that complement methods aimed at more indirect indicators such as plaque formation or genetic risk factors.

Nucleic acid-based blood liquid biopsies have been suggested as an attractive alternative due to their minimal invasiveness, scalability, and ability to capture cell and disease-specific molecular features^20–26^. Applications in oncology are currently being explored^27–30^. We have hypothesized that cfDNA methylation signatures, in particular, could bridge the gap between broad but nonspecific staging markers (e.g., NfL, GFAP) and narrow, disease-specific proteins (e.g., tau, α-synuclein), offering the specificity, sensitivity, and scalability needed for earlier detection and intervention^13^.

Impactful studies have demonstrated that methylation signatures can be leveraged to identify cellular origin across tissues^31^. Whole genome methylation profiles can be used to computationally deconvolute cfDNA fragments to their source tissues and have been applied to prenatal testing, oncology, and organ transplants^32^.

Subsequent studies have extended this concept with a more comprehensive human cell-type methylation atlas, revealing that circulating cfDNA encodes distinct cell-of-origin signatures in both healthy and diseased states^33^. For neurodegenerative disease, specifically, we, and other groups, have shown that elevated circulating levels of cfDNA from specific neurons and glia correlate with diseases like Alzheimer’s disease^34^, traumatic brain injury, and MS^35^.

However, these prior cfDNA methylation studies have relied on methods that include bisulfite conversion, PCR amplification, and short-read sequencing. While impactful in advancing the field, the approach introduces major limitations. Bisulfite- and PCR-based methods degrade DNA^36^ and introduce amplification bias^37^, limiting resolution and translational reliability.

In our pilot study, we detected elevated cortical neuron-derived cfDNA in the blood of AD and MCI patients compared to controls, using a targeted bisulfite/PCR assay at a single CpG site^34^. While this provided early proof of concept, it also underscored the limitations of locus-restricted, bisulfite- and PCR-based methods. The present study builds directly on that work, overcoming these constraints with native methylation profiling via nanopore sequencing. To enhance cortical neuron specificity, we expanded the methylation reference atlas to include additional neuronal and glial populations, which also revealed potential applications across other neurodegenerative diseases. Here, we establish proof of concept that cell type-resolved cfDNA deconvolution provides a scalable and clinically feasible framework for detecting and tracking neurodegeneration and differentiating neurodegenerative diseases.

## 2. METHODS

### 2.1 Sample Acquisition

#### 2.1.1 Purified Neural Cell Types

Purified human neurons and glial cells were obtained from commercial vendors (Accegen and Creative BioArray; Supplemental Table 1). Cell types included both primary and induced pluripotent stem cell (iPSC)-derived cortical and dopaminergic neurons, as well as primary astrocytes, Schwann cells, and microglia.

Postmortem human spinal cord tissue was procured through BioIVT under institutional protocols for deidentified human biospecimen research (BioIVT ASTERAND® Normal Human Tissue). Upon receipt, tissue was rapidly processed to preserve DNA integrity. Spinal motor neurons were isolated using a Percoll gradient method adapted from previous methods^38^, and optimized for mature human spinal cord tissue. Briefly, tissue was mechanically dissociated and layered onto dual isosmotic Percoll gradients (35% and 12.5%). Samples were centrifuged at 800 xg for 30 minutes, after which the 12.5% layer was collected for downstream processing. This fraction was enriched for spinal motor neurons and used for subsequent DNA extraction and methylation profiling. Optimization steps included adjusting gradient concentrations, centrifugation parameters, and handling conditions to account for the increased density and fragility of mature human neurons.

Genomic DNA extracted from these purified populations were used to generate whole genome nanopore methylation reference profiles for classifier training, evaluate cell type specificity, and validate the performance of methylation-based models for cfDNA deconvolution.

#### 2.1.1 Human Patient Blood Samples

A total of 219 human plasma samples (1 mL each) were obtained from BioIVT (Westbury, NY, USA), representing six clinically confirmed diagnostic categories: Alzheimer’s disease (AD; n = 45), mild cognitive impairment (MCI; n = 5), Parkinson’s disease (PD; n = 64), sporadic amyotrophic lateral sclerosis (SALS; n = 65), familial amyotrophic lateral sclerosis (FALS, n = 10), and cognitively normal, age-matched controls (HC, n = 30).

All samples were accompanied by case report forms (CRFs), informed consent documentation, and detailed clinical metadata. Demographic variables included age, sex, and ethnicity. Available clinical measures included Mini-Mental State Examination (MMSE) scores at multiple timepoints and cerebrospinal fluid (CSF) concentrations of disease-associated protein biomarkers. These included Aβ1-42, Aβ1-40, Aβ1-38, total tau, α-synuclein, and PARK7, quantified using validated commercial assays (Meso Scale Discovery [MSD], INNOTEST, and Covance platforms).

All BioIVT specimens were de-identified prior to receipt and used in accordance with institutional review protocols and ethical guidelines governing secondary use of human biological materials.

### 2.2 Sample Processing

All biospecimens were processed by Wasatch BioLabs (WBL), a high-throughput sequencing provider specializing in clinical and research applications of Oxford Nanopore Technologies sequencing. Sample processing followed standardized internal quality control protocols and was managed through a laboratory information management system (LIMS) to ensure traceability and data integrity.

#### 2.2.1 Genomic DNA

Genomic DNA (gDNA) was extracted from purified cell types using the DNeasy Blood & Tissue Kit (Cat # 69504) and libraries were prepared according to Wasatch Biolabs’ (WBL’s) Direct Whole Methylome Sequencing (dWMS) protocol, a modified whole genome ONT sequencing service optimized for methylation analysis. Extracted DNA was fluorometrically quantified using the Qubit dsDNA High Sensitivity Assay (Thermo Fisher Scientific) and assessed for purity and integrity using the NanoDrop spectrophotometer (Thermo Fisher Scientific) and Agilent TapeStation 4200 (Agilent Technologies), respectively.

Following gDNA dWMS sequencing, cell types included in model training achieved an average of 15x global coverage, ensuring robust methylation signal detection and accurate cell-type classification.

#### 2.2.2 Cell-Free DNA

cfDNA was extracted from human plasma samples using the MAGicBead™ cfDNA Isolation Kit (Zymo, Cat # D4086). cfDNA library preparation was performed according to WBL’s Direct Methylation Sequencing (dMS) for cfDNA service, an ONT-based cfDNA sequencing service for unbiased, native cfDNA sequencing. cfDNA quantity was measured using Qubit fluorometric quantitation (Thermo Fisher Scientific), and sample purity and integrity were assessed using NanoDrop spectrophotometry and the Agilent TapeStation 4200, respectively, as described above for gDNA.

### 2.3 Data Generation

DNA methylation array data for purified cortical neurons, dopaminergic neurons, and iPSC-derived spinal motor neurons were generated at the University of Utah using the Illumina Infinium MethylationEPIC v2 array platform in alignment with manufacturer protocols.

For nanopore sequencing, gDNA and cfDNA libraries were sequenced on the ONT PromethION platform. Raw signal data (POD5 files) were basecalled and aligned to the GRCh38 human reference genome using Dorado (https://github.com/nanoporetech/dorado), an open-source ONT basecaller. Methylation calls were extracted using ONT’s modkit software (https://github.com/nanoporetech/modkit), to produce per-read methylation summary files.

All methylation data from gDNA and cfDNA sequencing were converted to CH3, a proprietary file format developed by Wasatch BioLabs for downstream methylation analysis, as detailed below.

### 2.4 Differential Methylation Analysis

All array samples underwent standard preprocessing and normalization. Raw IDAT files were processed using the minfi R package. Beta values representing the fraction of methylation at each CpG site were obtained following SWAN (Subset-quantile Within Array Normalization). To assess data quality, beta value density plots were examined to confirm the expected bimodal distribution, characterized by peaks in the unmethylated (0.0–0.2) and methylated (0.8–1.0) ranges, and a flat valley in the intermediate methylation range. Samples failing to meet these distribution criteria were excluded from further analysis, and the remaining samples were re-normalized using the same procedures.

For all nanopore sequencing data, CH3 methylation files were created for each sample and processed with MethylSeqR, WBL’s open-source R package that that converts CH3 files into a queryable database, enables region/window summarization, differential methylation testing, and built-in QC plots (https://github.com/Wasatch-Biolabs-Bfx/MethylSeqR, v0.8.0). Methylation databases were indexed by individual CpG positions with the make_pos_db() function. Using the summarize_windows() function, these data were summarized into 200bp windows with a minimum of 5 CpGs. Differential methylation analyses were performed between each cell type and pooled human blood plasma using the calc_mod_diff() function. Differentially methylated regions (DMRs) were identified by collapsing windows using the collapse_windows() function, retaining regions with a minimum delta of at least 0.8 between cell type and blood plasma.

DMRs between each cell type and blood plasma were further filtered to remove overlapping DMRs between cell types. Resulting cell-specific DMRs were used in downstream classification to evaluate cell-type-specific cfDNA contributions in plasma from patients with neurodegenerative diseases.

### 2.5 Theoretical Model Validation

To evaluate the analytical performance of the cell type-specific classifiers, we generated *in silico* mixtures of cell type-specific cfDNA in a background of plasma-derived cfDNA.

Genomic DNA (gDNA) from each purified cell type was computationally fragmented to more closely mimic the size distribution of circulating cfDNA (see Supplemental Figure 1)^39^. This step ensured that cell-derived gDNA reads would be more comparable in size and structure to those found in patient-derived cfDNA samples.

Using these computationally fragmented reads, a series of *in silico* dilution datasets was constructed. Each dataset contained approximately 1,000,000 total reads and consisted of varying proportions of cell-specific reads spiked into a background of healthy donor plasma cfDNA. The dilution series included nine fractional compositions: 0%, 0.1%, 1%, 10%, 15%, 25%, 50%, 75%, and 100% cell-type-specific reads, with the remaining balance composed of plasma reads. This range was designed to reflect physiologically plausible concentrations of cell-free DNA originating from neuronal and glial sources. For each dilution point, cell-type-specific DMR classifiers were applied to the synthetic mixtures. Classifier scores were calculated for individual reads based on methylation concordance with the reference DMR set corresponding to the target cell type. Predicted cell fractions were derived by aggregating classified reads and were then compared to the known spiked-in proportions.

Classifier performance was assessed by calculating the coefficient of determination (R^2^) between predicted and actual cell fractions. Linearity and sensitivity were evaluated across the dilution range, and classifier specificity was tested by applying each model to unrelated cell types. Additional comparisons were performed using DMRs derived from iPSC-based models to assess the impact of training source on classification fidelity.

This theoretical validation enabled benchmarking of model accuracy and demonstrated the potential of the classifier framework to detect low-frequency cfDNA signals in a complex plasma background.

### 2.6 Clinical Analyses

As an initial exploratory step, the distribution of cell-type specific cfDNA classifications was visualized across diagnostic categories. For each cell type of interest (e.g., cortical, dopaminergic, or spinal motor neurons), boxplots displaying the percentage of reads classified to that cell type across different disease states were generated. These plots were overlaid with jittered individual points to highlight within-group variability and potential diagnostic separation. This visualization provided early evidence of cell-type specific cfDNA differences between disease and control groups, motivating subsequent classification modeling.

To quantitatively assess the diagnostic value of these cell-type specific cfDNA signals, a logistic regression classification framework was developed using a stratified train-test split. Only samples from the disease group(s) of interest and comparator group(s) were included. Binary labels were assigned accordingly. The dataset was then partitioned into a training set (70%) and a testing set (30%) using stratified sampling to maintain class balance.

The training set was used to fit a logistic regression model with one or more specified predictors (e.g., % spinal-classified reads) corresponding to different neuronal or glial populations. Importantly, the training set was also used to identify the optimal decision threshold for classification, selected based on Youden’s J statistic derived from the receiver operator characteristic (ROC) curve. This threshold was fixed and carried forward to the test set without modification, and is reported here as “optimal threshold”.

All performance metrics, including sensitivity, specificity, accuracy, and area under the ROC curve (AUC) with 95% confidence intervals were evaluated exclusively on the test dataset to avoid overfitting and ensure generalizability. ROC curves with confidence intervals and confusion matrix heatmaps were generated to visualize model performance. All statistical analysis and figure generation were performed in R using the caret, pROC, dplyr, tibble, and ggplot2 packages.

### 2.7 Classifier Performance and Statistical Analysis

All analyses were performed using R (v4.3.0) and Python (v3.10), supported by in-house developed scripts. Statistical analyses were performed to evaluate the quantitative accuracy and diagnostic performance of cfDNA methylation classifiers across both computational and clinical datasets. For analytical validation using *in silico* dilution series, predicted cfDNA fractions were compared to known input values, and model performance was quantified using linear regression and R^2^. These analyses assessed the linearity and concordance of predicted cell fractions across a wide dynamic range of cell-type-specific input levels.

To assess differences in cell classification signals between disease groups, we employed the Wilcoxon rank-sum test (Mann–Whitney U test). This non-parametric statistical test was selected due to its robustness against violations of normality and its suitability for comparing distributions of continuous variables between two independent groups. Specifically, we compared cell-type-specific classification scores (e.g., cortical or dopaminergic cfDNA percentages) between disease states (e.g., Parkinson’s disease vs. age-matched controls, or ALS vs. Alzheimer’s disease). All p-values obtained from pairwise Wilcoxon tests were subsequently adjusted for multiple comparisons using the Benjamini-Hochberg procedure to control the false discovery rate (FDR). Adjusted p-values < 0.05 were considered statistically significant. These analyses were conducted in R (v4.5.1) using the wilcox.test() function from base R and p.adjust() for FDR correction.

The same statistical framework was applied to ancillary analyses of protein-based biomarkers, including amyloid-β isoforms, total tau, and Mini-Mental State Examination (MMSE) scores. These data were plotted as boxplots based on raw concentration values or scores provided by the sample supplier (BioIVT). No normalization, transformation, or additional statistical testing was performed, as we did not have access to the proprietary diagnostic pipelines used by clinical providers. Accordingly, these results are presented solely for qualitative context and visual comparison with cfDNA-derived signals (Supplemental Figure 11).

For classification tasks in the clinical cohorts, diagnostic performance was assessed through ROC curve analysis. Classifier scores for each sample were generated based on read-level concordance with cell type-specific DMRs, and binary classifications were evaluated for their ability to distinguish between disease and control groups, as well as between other disease groups. The following performance metrics were calculated: AUC, 95% confidence intervals for AUC (lower and upper bounds), sensitivity, specificity, positive predictive value (PPV), negative predictive value (NPV), overall accuracy, balanced accuracy, and the optimal classification threshold, which was determined using Youden’s J statistic. These metrics were computed for each classifier individually, including cortical neuron-, dopaminergic-, and spinal motor neuron-derived cfDNA, as well as for a combined model incorporating all cell types.

## 3. RESULTS

### 3.1 Whole-genome nanopore profiling identifies cell type-specific methylation signatures

To establish our methylation reference atlas for cfDNA classification, we performed whole-genome methylation sequencing on genomic DNA from pooled human blood plasma and eight primary neuron and glial cell types using Wasatch BioLabs’ (WBL) Direct Whole Methylome Sequencing (dWMS) platform. This Oxford Nanopore Technologies (ONT)-based method captured native 5mC/5hmC methylation with single-molecule and CpG resolution.

Differential methylation analyses between plasma and each neuronal and glial cell type identified millions of differentially methylated CpGs for each comparison using a threshold of Δβ ≥ 0.8 (Supplemental Table 2). Among the primary neurons, cortical neurons exhibited the highest number of differentially methylated CpG sites (4.28 million), followed by dopaminergic (1.75 million) and spinal motor neurons (1.54 million). Differential methylation analysis of glial cells—astrocytes, microglia, and Schwann cells—compared to pooled blood plasma yielded between 1.24 and 1.43 million differentially methylated CpG sites (Supplemental Table 2). Differentially methylated regions (DMRs) identified with nanopore sequencing drastically outnumbered DMRs identified via array (Supplemental Table 2).

To assess whether induced pluripotent stem cell (iPSC)-derived neurons could serve as a more accessible alternative to primary cells for classifier development, we included iPSC-derived cortical, dopaminergic, and spinal motor neurons in our analyses. These cell types yielded a comparable number of differentially methylated CpG sites relative to their primary counterparts (data not shown).

To assess cell type specificity, DMRs were compared across all neural populations, including both primary and iPSC-derived cell types. Thousands of overlapping and unique DMRs were identified per cell type. Figure 1A reports total, overlapping, and unique DMRs, with directional visualizations of methylation change (hypermethylation in red, hypomethylation in blue). Primary cortical neurons displayed 117,695 unique DMRs, primary dopaminergic neurons displayed 29,086 unique DMRs, primary spinal motor neurons displayed 89,648 unique DMRs (Figure 1A). Unique, non-redundant, cell type-specific DMRs for these cell types were used for downstream classifier development.

**Figure 1.**
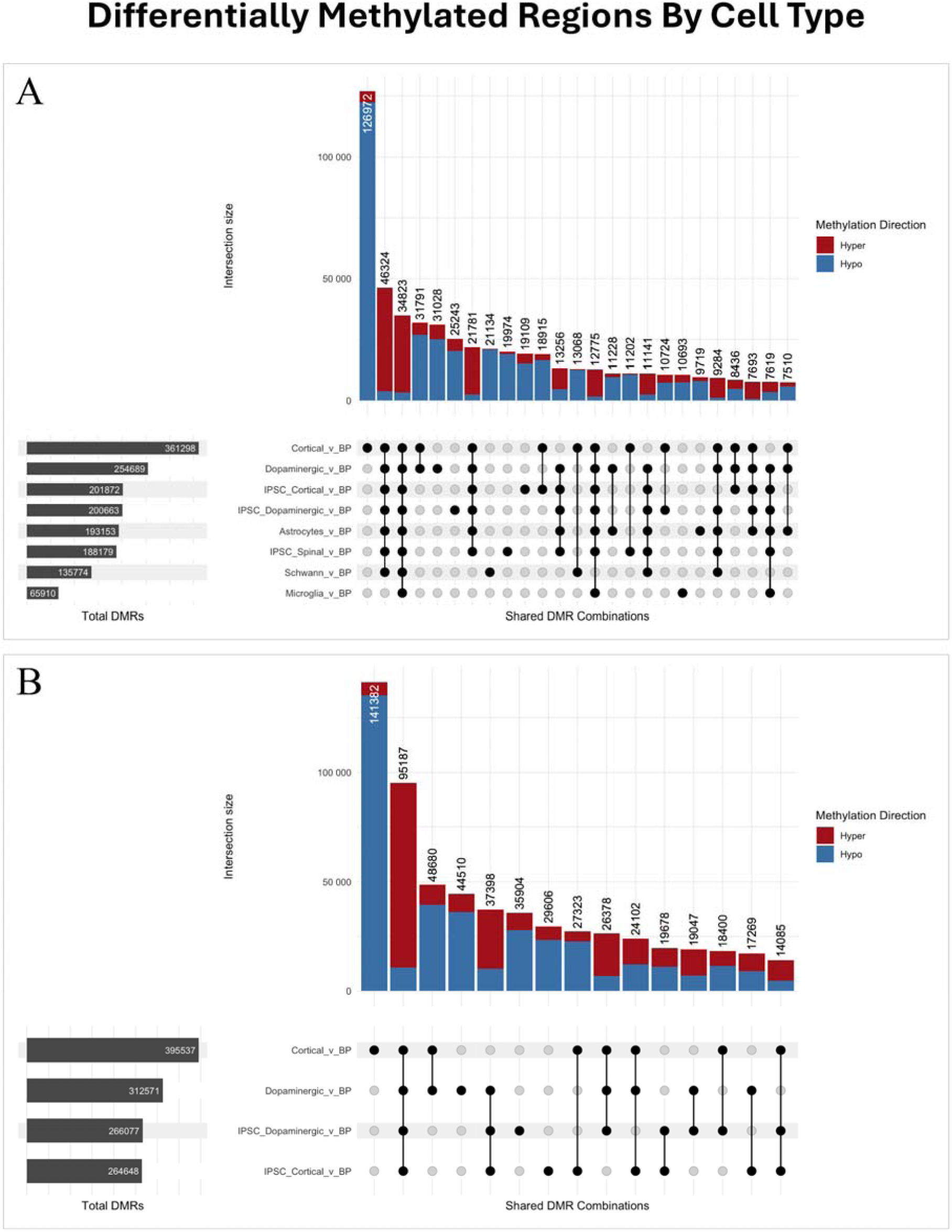
(A) UpsetR plot showing cell-specific DMRs and overlap between each cell type. (B) UpsetR plot showing cell specific DMRs and overlap between ISPC’s and primary cells. Both panels include hypo and hypermethylation annotations to explore epigenetic commonalities and differences among diseases.

Comparing DMRs between iPSC-derived neurons and blood plasma to their primary counterparts revealed substantial divergence. Only 28.3% of DMRs in iPSC-derived cortical neurons overlapped with primary cortical neurons, 38.1% with dopaminergic neurons, and 6.8% with spinal motor neurons (Figure 1B). Although the total number of DMRs was comparable, their content and genomic distribution differed widely.

### 3.2 *In silico* validation demonstrates high analytical accuracy and specificity of cfDNA classifiers

To assess the analytical accuracy and specificity of our DMR-based classifiers, we generated *in silico* cfDNA-like gDNA mixtures containing defined proportions of cell type-specific reads in a plasma background and applied our classifiers. To better approximate native cfDNA properties, we computationally fragmented genomic DNA reads from purified cells, which were initially longer than plasma-derived cfDNA. Resultant fragment distributions more closely resembled circulating cfDNA (Supplemental Figure 1).

The cortical neuron classifier displayed the highest predictive accuracy, with a near-perfect correlation between predicted and actual cell fractions across the full dilution series (R^2^ to identity = 0.998; Figure 2A). Similarly high performance was observed for primary dopaminergic neurons (R^2^ to identity = 0.993, Figure 2B), astrocytes (R^2^ = 0.993, Figure 2D), Schwann cells (R^2^ to identity = 0.986, Figure 2E), and microglia (R^2^ to identity = 0.992, Figure 2F), each achieving R^2^ to identity values above 0.98.

**Figure 2.**
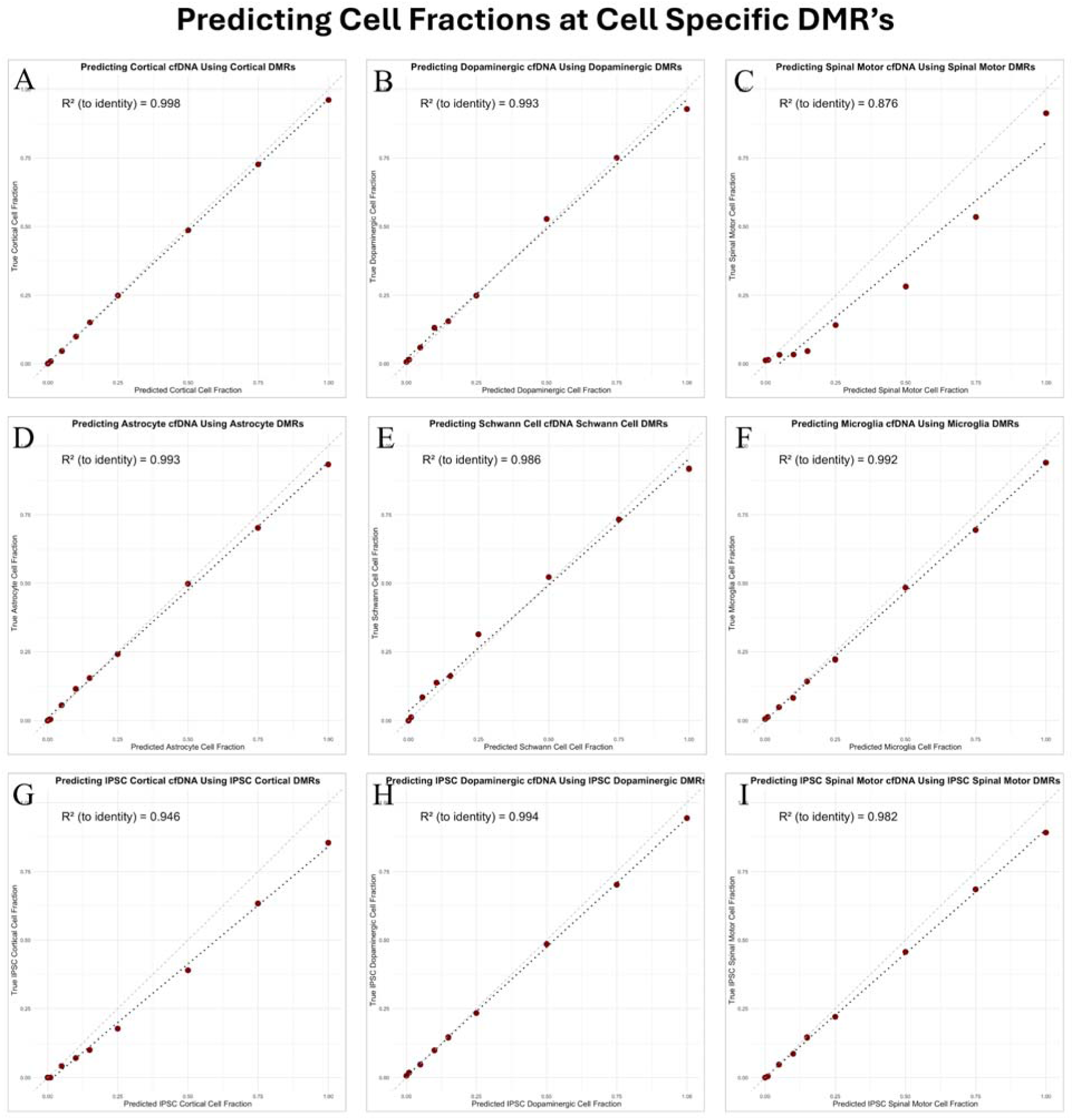
Correlation plots comparing predicted versus actual cell-type proportions in synthetic dilution datasets for each cell type. (A) Predicting the proportion of cortical derived cfDNA using cortical specific DMRs. (B) Predicting the proportion of dopaminergic neuron derived cfDNA using dopaminergic neuron specific DMRs. (C) Predicting the proportion of spinal motor neuron derived cfDNA using dopaminergic neuron specific DMRs. (D) Predicting the proportion of astrocyte derived cfDNA using astrocyte specific DMRs. (E) Predicting the proportion of schwann cell derived cfDNA using schwann cell specific DMRs. (F) Predicting the proportion of microglial derived cfDNA using microglial specific DMRs. (G) Predicting the proportion of ipsc cortical neuron derived cfDNA using ipsc cortical neuron specific DMRs. (H) Predicting the proportion of ipsc dopaminergic neuron derived cfDNA using ipsc dopaminergic neuron specific DMRs. (I) Predicting the proportion of ipsc spinal motor neuron derived cfDNA using ipsc spinal motor neuron specific DMRs.

Strong linearity was also observed across iPSC-derived *in silico* mixtures, including iPSC-derived cortical neurons (R^2^ to identity = 0.946, Figure 2G), dopaminergic neurons (R^2^ to identity = 0.994), and spinal motor neurons (R^2^ to identity = 0.982, Figure 2I). In contrast, primary spinal motor neuron-derived fractions and models developed from iPSC-derived neurons exhibited substantially reduced performance (R^2^ to identity = 0.876).

To assess cell type specificity, we also applied each classifier to *in silico* datasets from other cell types. In all cases, slopes for the other cell types was near zero, indicating high model specificity for each cell type (Supplemental Figures 2-10).

### 3.3 Clinical proof of concept for cfDNA classifiers in patient plasma

#### 3.3.1 Neuron-specific cfDNA elevations distinguish Alzheimer’s, Parkinson’s, and ALS samples from controls

After establishing the predictive ability of the models, we assessed their clinical potential by applying the primary neuron classifiers to 219 plasma samples from healthy controls and patients with MCI, AD, PD, or ALS. This analysis revealed cfDNA elevations reflecting targeted neuronal cell loss across diseases, underscoring the diagnostic potential of neuron-specific cfDNA methylation signals (Figure 3A–F).

**Figure 3.**
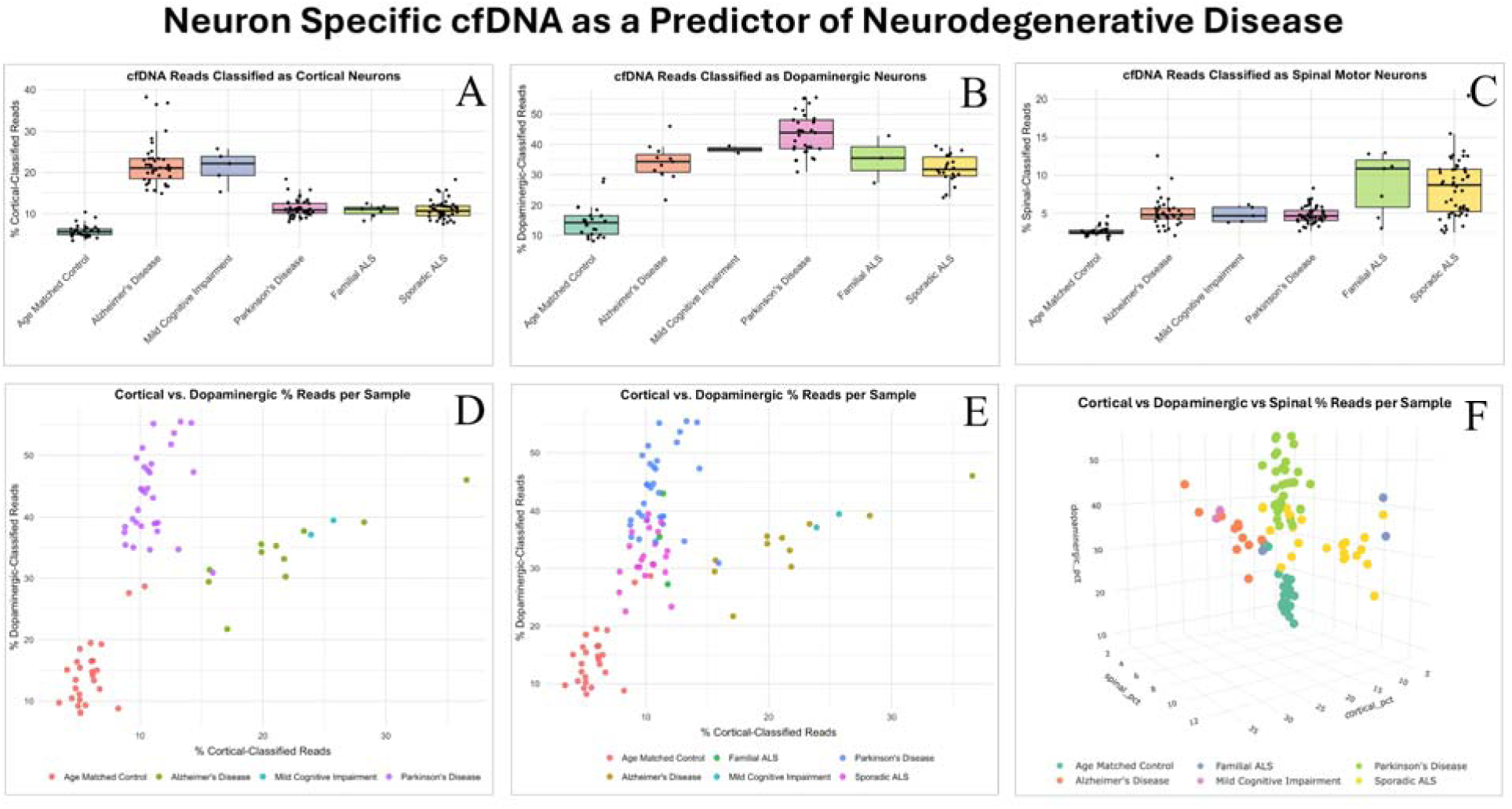
Visualization of neuron-specific cfDNA classifications across patient groups. (a), (b), and (c) show the proportion of cortical, dopaminergic, and spinal cfDNA, respectively, across AD, MCI, PD, ALS, and control samples. (d) and (e) plot cortical vs. dopaminergic cfDNA percentages in a bivariate space to illustrate disease separation with and without ALS patients. (f) plots cortical vs. dopaminergic vs. spinal in a 3d model to illustrate disease separation.

Cortical neuron-derived cfDNA was significantly elevated in AD and MCI (22.19%, SD = 5.45%) when compared to controls alone (5.73%, SD = 1.53%; p = 1.54e-28), and when compared to controls, PD, and ALS combined (10.48%, SD = 4.40%; p = 1.08e-21; Figure 3A, D). Dopaminergic neuron-derived cfDNA was significantly elevated in PD (42.60%, SD = 10.60%) when compared to controls alone (15.11%, SD = 5.19%; p = 6.92e-12; Figure 3B, E), and when compared to AD, MCI, ALS, and controls combined (29.63%, SD = 12.12%; p = 1.00e-12). Spinal motor neuron-derived cfDNA was significantly elevated in familial and sporadic ALS (7.80%, SD = 4.45%) when compared to controls alone (4.61%, SD = 2.11%; p = 7.05e-08) and when compared to all other groups (4.61%, SD = 2.11; p = 7.05e-8; Figure 3C, F).

By contrast, classifiers for astrocytes, Schwann cells, microglia, and iPSC-derived neurons showed no consistent disease-associated changes in patient plasma (Supplemental Figure 12). Astrocyte and microglial cfDNA remained stable across all groups, and while Schwann cell cfDNA showed a modest increase in ALS relative to controls, this effect was not significant when other diseases were included. Likewise, iPSC-derived neuron classifiers failed to distinguish relevant disease states from controls.

Together, these results are consistent with our broader observation that iPSC-derived neuron methylation profiles differ substantially from those of primary neurons. Importantly, they provide further support that the cfDNA signals we detected reflect true cell-of-origin specificity, as cfDNA derived from distinct neuronal subtypes aligned closely with the selectively vulnerable populations in AD, PD, and ALS. These findings reinforce the diagnostic potential of cell type-resolved cfDNA profiling while also underscoring that iPSC-derived neurons, although accessible, cannot accurately recapitulate primary *in vivo* methylomes required for cell of origin classifiers.

#### 3.3.2 Integrating multiple neuronal cfDNA signals enhances disease prediction

To further improve disease prediction accuracy, we applied cortical, dopaminergic, and spinal motor neuron classifiers as an integrated model combining all three. The integrated model was tested in both binary (e.g., AD vs. controls, PD vs. controls, ALS vs. controls) and multi-disease (e.g., AD vs. all other diseases and controls) contexts via ROC analysis, which enabled side-by-side comparisons of univariate and multivariate models.

The multivariate classifiers yielded comparable or higher AUC values than the univariate counterparts, generally displaying improved sensitivity, specificity, and overall accuracy. For the binary AD vs. control analysis, the individual cortical neuron classifier produced an AUC = 1.0000 (95% CI: 1.0000–1.0000), with 100% sensitivity, 100% specificity, corresponding positive and negative predictive values (PPV and NPV) of 1.0000, and balanced accuracy of 100% at the optimal classification threshold of 0.5 (Figure 4). The multi-cell model that integrated all 3 neurons maintained perfect performance, with AUC = 1.0000 (95% CI: 1.0000–1.0000), with 100% sensitivity, 100% specificity, corresponding positive and negative predictive values (PPV and NPV) of 1.0000, and balanced accuracy of 100% at the optimal classification threshold of 0.5.

**Figure 4.**
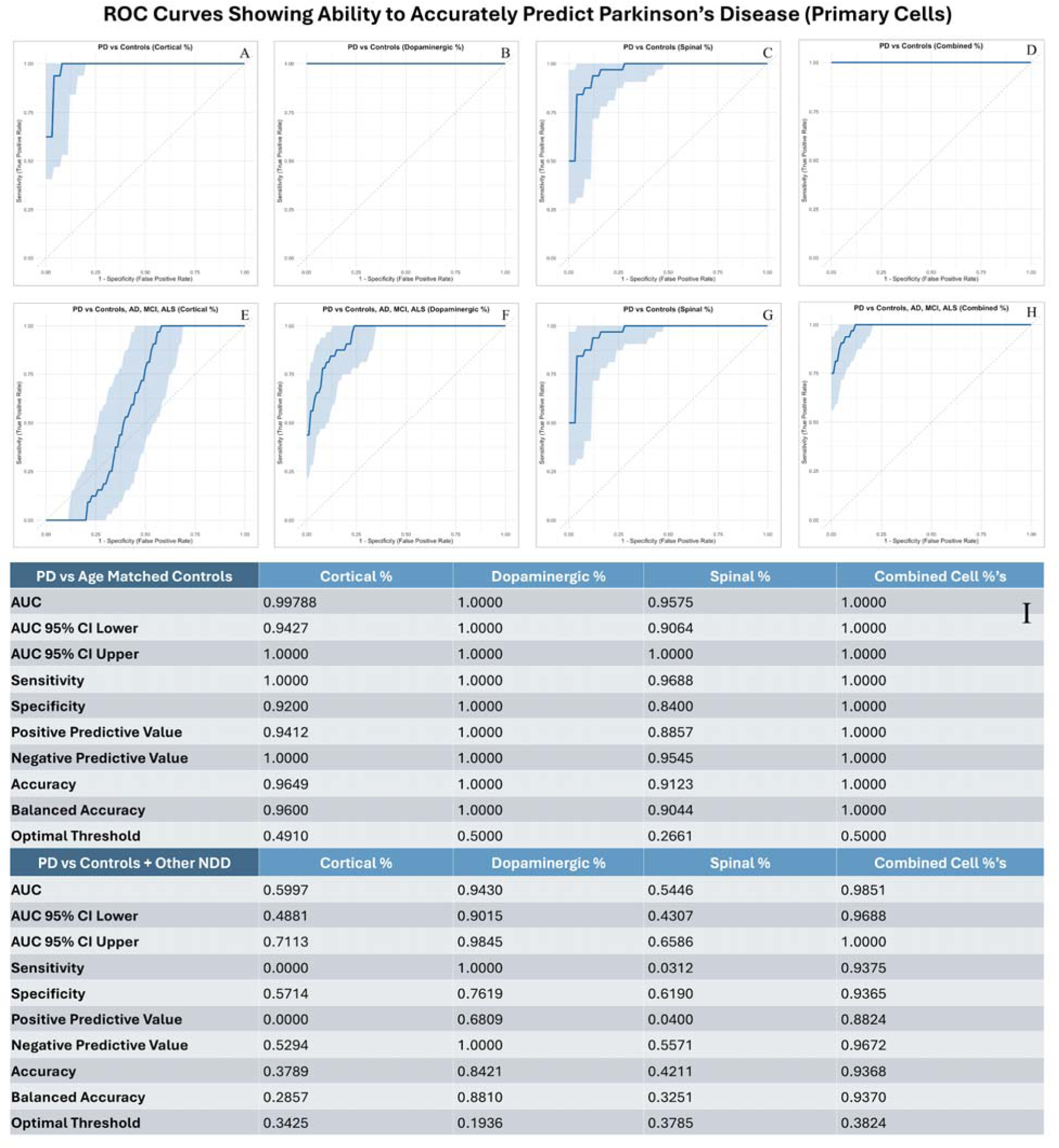
Receiver Operating Characteristic (ROC) curves for predicting Alzheimer’s Disease using Primary Cell derived models for (a)(e) cortical, (b)(f) dopaminergic, and (c)(g) spinal neuron-derived cfDNA, alone and in (d)(h) combination. (i) shows the respective stats associated with each panel. Panels compare classification performance against both controls and other neurodegenerative diseases, showcasing the additive value of combining multiple cell type signals.

To further demonstrate the added value of integrating multiple cell-type signatures into one model, we compared the performance of the combined classifier to the corresponding univariate models across PD and ALS cases. In the PD comparisons (Figure 5), while both the dopaminergic and combined models achieved perfect separation from controls (AUC = 1.0000), only the combined model maintained strong performance when distinguishing PD from ALS and AD. In contrast, cortical and spinal motor classifiers, which performed well in PD vs. control settings (AUC = 0.9979 and 0.9575, respectively), showed steep declines in the three-class setting (AUC = 0.5997 and 0.5446). The combined model effectively aggregated discriminative signals across relevant and partially overlapping cell types, preserving high accuracy across a more complex clinical landscape.

**Figure 5.**
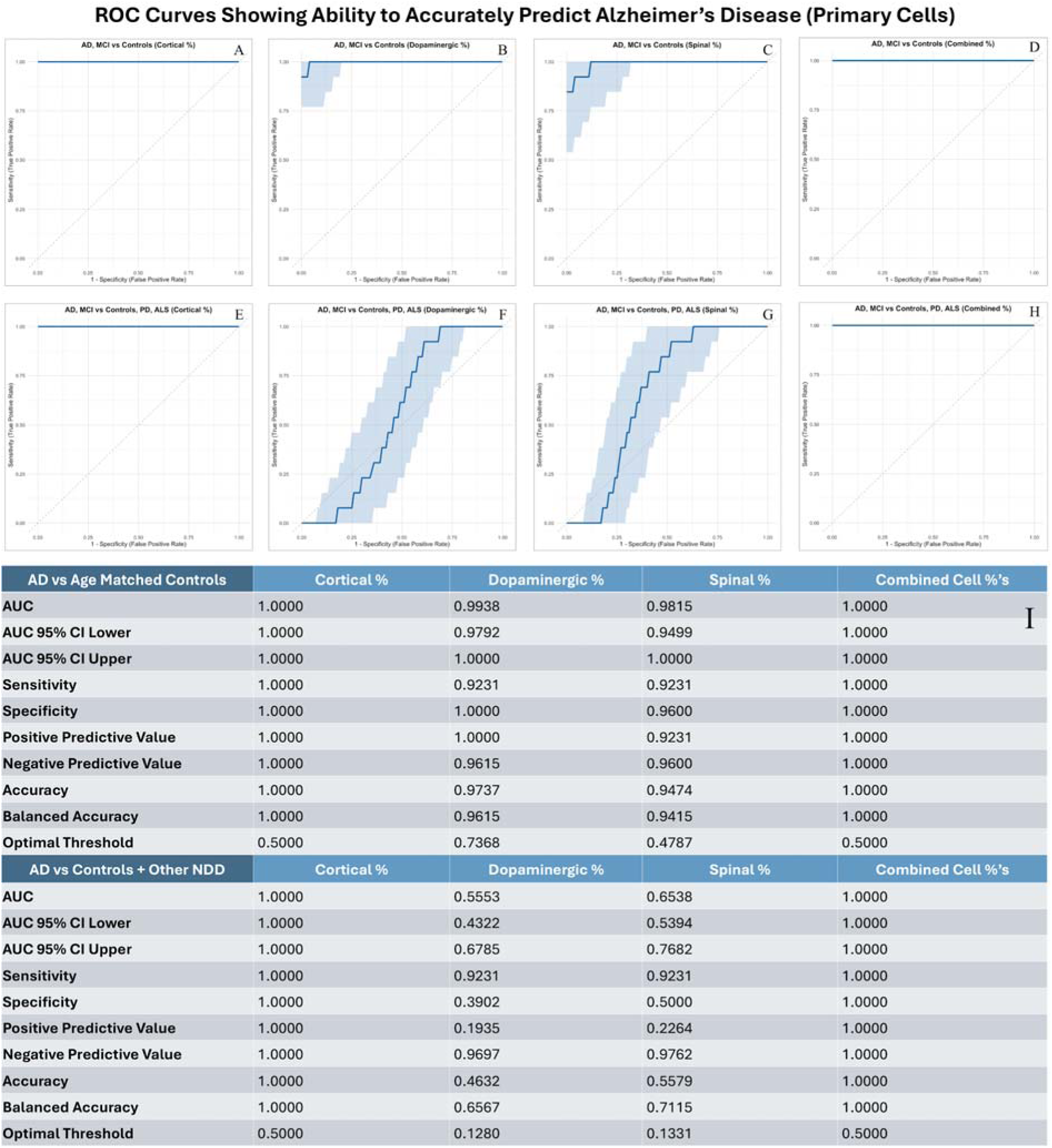
Receiver Operating Characteristic (ROC) curves for predicting Parkinson’s Disease using Primary Cell derived models for (a)(e) cortical, (b)(f) dopaminergic, and (c)(g) spinal neuron-derived cfDNA, alone and in (d)(h) combination. (i) shows the respective stats associated with each panel. Panels compare classification performance against both controls and other neurodegenerative diseases, showcasing the additive value of combining multiple cell type signals.

A similar pattern held for ALS (Figure 6). The spinal motor classifier alone yielded strong binary classification (AUC = 0.9680), but its specificity dropped in multi-disease comparisons (AUC = 0.6538). Other univariate classifiers, while showing high AUCs in ALS vs. control settings (cortical: 0.9696; dopaminergic: 0.9872), again lacked discriminative power across multiple diseases (AUCs = 0.5600 and 0.5497). By contrast, the combined ALS model sustained high discrimination (AUC = .9905), highlighting its ability to integrate weak but complementary signals from across cell types to improve disease specificity.

**Figure 6.**
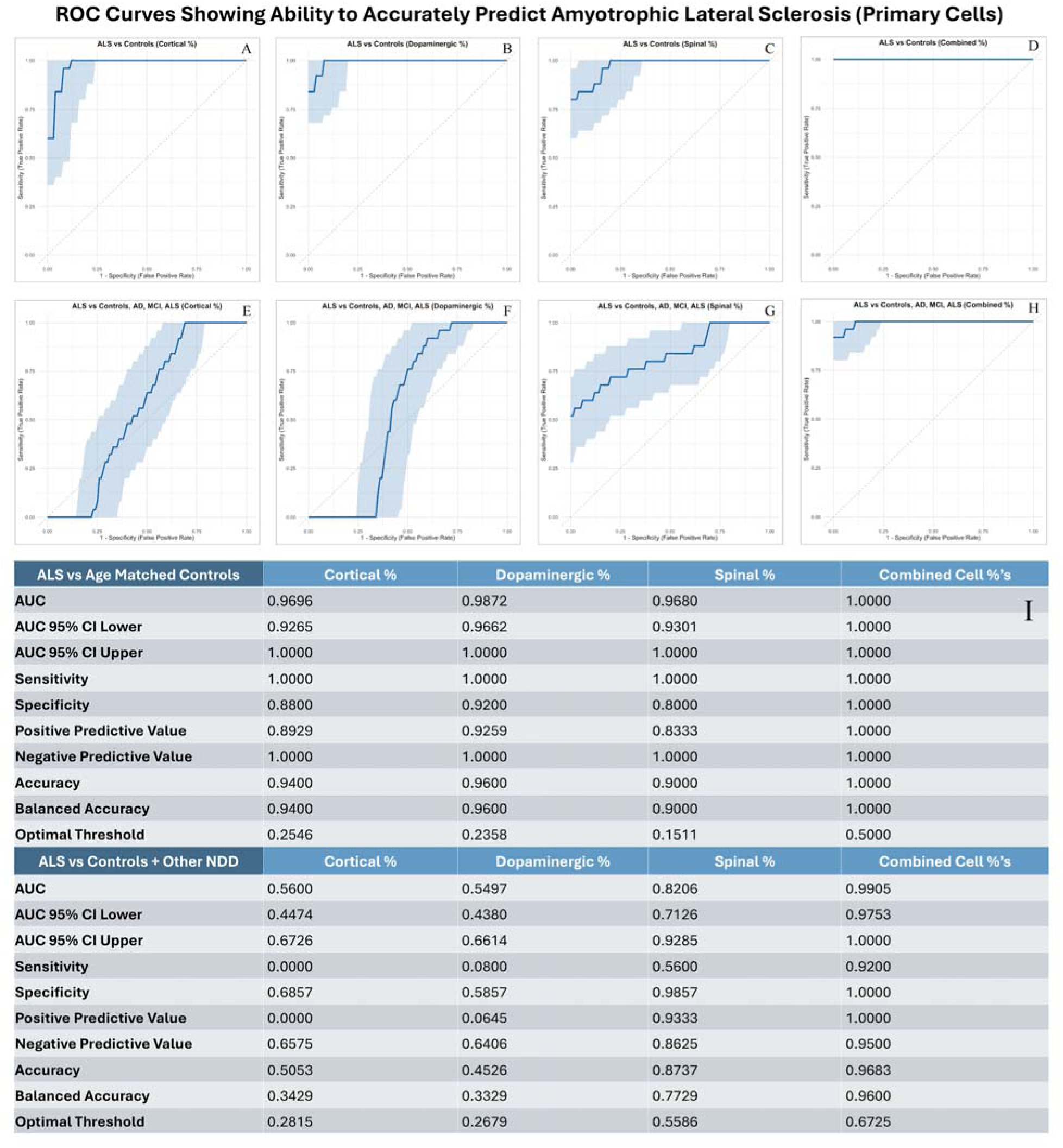
Receiver Operating Characteristic (ROC) curves for predicting Amyotrophic Lateral Sclerosis using Primary Cell derived models for (a)(e) cortical, (b)(f) dopaminergic, and (c)(g) spinal neuron-derived cfDNA, alone and in (d)(h) combination. Panels compare classification performance against both controls and other neurodegenerative diseases, showcasing the additive value of combining multiple cell type signals.

These cell type-specific disease predictions were completely lost when iPSC-derived models were applied, whether individually or in combined multi-cell formats, and in both binary and multi-disease contexts (Supplemental Figures 13–15). Together, these results demonstrate that while neuron-specific classifiers capture relevant disease signals, integrating cfDNA methylation features across neuronal populations provides greater accuracy and specificity for predicting disease in real-world contexts where all disease diagnoses are possible.

## 4. DISCUSSION

This study establishes a scalable liquid biopsy framework for cell type-resolved detection of neurodegeneration from cfDNA methylation profiling. To address the limitations of our prior Alzheimer’s work^34^, we generated a whole-genome nanopore methylation atlas of six primary neural cell types and developed classifiers that sensitively and specifically detect cortical, dopaminergic, and spinal motor neuron DNA. Applying these classifiers to blood plasma from patients with AD, PD, and ALS, we found that cfDNA from vulnerable neuronal populations can be detected in circulation: cortical cfDNA was elevated in AD, dopaminergic cfDNA in PD, and spinal motor neuron cfDNA in ALS, achieving an AUC > 0.98 (Figures 4–6). These signals were disease-associated and orthogonal, enabling clear separation of AD, PD, and ALS in multiclass comparisons, and distinction of cell type-specific pathologies rather than generic neurodegeneration.

Integrating methylation signatures from multiple neuronal populations into a multivariate model improved sensitivity, specificity, and overall accuracy compared to individual classifiers, highlighting the value of capturing the broader landscape of neurodegeneration. Together, these findings suggest potential utility of cfDNA methylation analysis in detecting and monitoring cell type-specific neurodegeneration. Applied in concert with other biomarkers, cfDNA methylation could enhance the molecular precision of disease prediction.

Our approach builds on established workflows for cfDNA cell-of-origin deconvolution pioneered in oncology, organ transplantation, and prenatal testing. These applications rely on the observation that DNA methylation signatures are highly cell type-specific, reproducible, and stable. Landmark studies have shown that methylation patterns are >99% stable across 39 sorted human cell types derived from 205 healthy tissues, and that replicates of the same cell type are >99.5% identical^31^. These findings emphasize that methylation-based identity programs are robust to environmental perturbation and therefore provide a biologically reliable substrate for cfDNA deconvolution^30,32,33^, and demonstrates that cfDNA carries rich, real-time information about tissue-of-origin injury and cellular turnover.

While neurological applications of cfDNA deconvolution have lagged behind oncology^27,40^, early studies offer compelling evidence of its utility. These studies demonstrate that neuron-derived cfDNA can be detected in circulation and suggest concentrations correlate with clinical phenotypes in traumatic brain injury and multiple sclerosis^35^. More recently, our own pilot study demonstrated elevated cortical neuron-derived cfDNA in patients with AD and MCI compared to healthy controls^34^.

While these and other studies established early feasibility, they have also exposed limitations. Most prior efforts have relied on bisulfite conversion-based sequencing that targets limited CpG loci or on array-based bisulfite methods that interrogate only a fraction of the methylome at predefined sites. These approaches degrade DNA^36,41^, and introduce PCR biases^37^. Collectively, these constraints have historically reduced both the biological specificity and the translational utility of cfDNA methylation profiling.

Building on earlier work, our study overcomes these barriers by applying whole-genome, native nanopore sequencing. The human genome contains ~30 million CpG sites^31,42^, yet Illumina’s EPIC array only surveys ~900,000 (~3%), primarily within CpG islands and promoters^43^. While arrays have expedited methylation breakthroughs and advanced the field through standardized workflows, they systematically exclude ~97% of the methylome and disproportionately emphasize well-characterized regulatory regions.

In our prior work, we identified a cortical neuron cfDNA marker using array data^34^, but could not establish whether it was uniquely cortical, pan-neuronal, or coincident. Here, whole-genome nanopore sequencing of purified primary cortical neurons and human blood plasma captured the entire CpG context, revealing millions of differentially methylated sites compared to the thousands by EPIC array, a lower-than-expected discovery rate given probe coverage (Supplemental Table 2). Indeed, the array captured only ~0.215% of the DMRs identified between cortical neurons and plasma cfDNA, compared to the expected 3%. This highlights the disproportionate representation of regulatory hotspots in array-based studies and underscores the value of native sequencing for unbiased discovery of cell type-specific signatures.

We expanded the biological scope of our reference atlas—encompassing cortical, dopaminergic, and spinal motor neurons as well as glial populations—which further improved disease specificity (Figures 4–6). This expansion enabled us to eliminate overlapping DMRs and focus on regions that were uniquely cell type-specific relative to the other populations. When applied to human clinical plasma samples, multivariate models from the broader atlas resolved cases that were ambiguous or misclassified by single-cell classifiers (Figures 4-6), underscoring the additive value of combining complementary cell type-specific signals. This highlights both the strengths of native-read sequencing and the value of expanding the reference atlas, which improved biological resolution and enhanced the accuracy of disease classification.

We acknowledge that true cell type specificity requires progressively eliminating overlapping methylation signals by incorporating additional reference cell types. In principle, a fully comprehensive atlas would require coverage of the >200 histologically defined and >6,000 transcriptomically defined human cell types^45,46^. Such exhaustive representation is not currently feasible, but for the purposes of this study, we determined that our selected set of neural populations was sufficient to generate robust, cell type-specific classifiers, a conclusion supported by our results. Moving forward, we view this limitation as an opportunity. Our data suggest that expanding the atlas to include additional CNS and peripheral cell types will likely enhance analytical accuracy and broaden the clinical utility of the cfDNA classifiers for detecting and differentiating neurodegeneration in disease. However, the acquisition of primary neuronal and glial cell types is often logistically difficult due to the limited availability of human donor material.

We encountered this limitation when sourcing primary human spinal motor neurons, which are rarely available from commercial vendors. To address this, we directly isolated neurons from postmortem spinal cord tissue using an optimized Percoll gradient approach. Although this method was likely less specific than FACS-based sorting used in prior atlases, the selective elevation of spinal motor neuron cfDNA in ALS suggests that the core disease-relevant signatures were captured (Figure 3C, Figure 6). As an exploratory alternative, we evaluated iPSC-derived neurons as a more accessible substitute. However, these cells failed to perform in clinical testing (Supplemental Figures 8–10), consistent with our qualitative observation that iPSC-derived methylomes differ substantially from their primary counterparts. Together, these findings underscore that primary tissue-derived references are critical for developing robust, biologically relevant classifiers.

Several other limitations of the present study should be noted. Single-time-point blood plasma samples are inherently retrospective and cross-sectional, limiting the ability to assess temporal cfDNA dynamics. Longitudinal studies will be essential to determine whether cfDNA changes can detect neurodegeneration before clinical symptoms emerge and to evaluate how reliably they track progression and therapeutic response. Notably, in our cohort, all individuals with MCI who subsequently progressed to AD exhibited elevated cortical cfDNA at the time of sampling. While this suggests that neuron-derived cfDNA can capture disease-relevant signals before clinical disease conversion, direct longitudinal AD studies will be required to confirm this observation. In addition, sample sizes of rarer diagnostic subgroups, such as MCI and familial ALS, were small and lacked demographic diversity, highlighting the need for larger, more representative cohorts to validate population-level generalizability.

We acknowledge that other neuronal and glial cell types with clear diagnostic relevance, such as oligodendrocytes (multiple sclerosis), remain absent from our atlas due to sourcing challenges. Incorporating these populations, along with additional peripheral cell types, will be important for refining background correction, improving disease-specific resolution, and broadening clinical applicability. Nevertheless, the robust detection of highly specific cfDNA signals across hundreds of patient plasma samples underscores the strength of our current framework and highlights its potential for translational impact even at this stage of development.

Unlike static biomarkers such as protein concentrations or imaging, which often change slowly and variably, cfDNA has a short half-life of minutes to hours^19,49^, providing a near real-time molecular snapshot of neuronal injury. In our cohort, we also explored the protein biomarker data that accompanied the patient plasma samples (Supplemental Figure 11). However, because only raw concentrations were available and we did not have access to the proprietary diagnostic pipelines used by providers, these analyses were considered ancillary and not central to our conclusions. We, instead, focused on cfDNA methylation signals as the primary endpoint in alignment with clinically-verified diagnoses, while recognizing that protein biomarkers remain an important complementary modality that will be explored more explicitly in the future.

This dynamic nature makes cfDNA a particularly attractive candidate for monitoring disease progression and therapeutic responses, offering sensitivity to short-term molecular changes that may not be captured by traditional approaches. In clinical practice, scalable cfDNA assays could support earlier diagnosis, presymptomatic risk stratification, and real-time monitoring of disease activity. In therapeutic development, cfDNA classifiers could accelerate clinical trials by improving patient selection and providing dynamic, objective endpoints, while preclinical applications may extend to animal models to track cell type-specific degeneration *in vivo*.

Our findings demonstrate that cfDNA methylation profiling can capture selective neuronal cell death with high specificity, establishing a biologically grounded and technically scalable framework for noninvasive detection and differentiation of neurodegenerative diseases. Future work expanding the reference atlas, incorporating additional neuronal and peripheral cell types, and conducting longitudinal studies will be critical to establish clinical validity. With these refinements, cfDNA classifiers could improve diagnostic timelines and enable earlier intervention across a spectrum of conditions, including AD, PD, and ALS. Beyond these diseases, the modular architecture of independent but combinable classifiers provides a flexible platform that could be extended to other neurological disorders with selective cellular vulnerabilities, such as multiple sclerosis, frontotemporal dementia, and cerebellar ataxias^16,50–52^. More broadly, this work supports cfDNA methylation profiling as a versatile biomarker platform adaptable to both research and clinical contexts.

## Data Availability

All data produced in the present study are available upon reasonable request to the authors

## ACKNOWLEDGEMENTS/CONFLICTS/FUNDING SOURCES

This research was supported by two primary sources: (1) a Sponsored Research Agreement between Brigham Young University (BYU) and Renew Biotechnologies, LLC, which wholly owns Resonant, LLC, and (2) Research Catalyst Program funding from the Alzheimer’s Disease and Dementia Research Center (ADRC) at Utah State University (USU). All authors are affiliated with Renew Biotechnologies and its subsidiary, Resonant, LLC, both for-profit entities, representing potential financial conflicts of interest. All work was conducted in accordance with institutional policies on conflict of interest disclosure and management.

