## Supplemental Material for "Circulating Cell-Free DNA Methylation Profiles Enable Disease-Specific Detection of Alzheimer’s, Parkinson’s, and ALS from Blood"

### 7. SUPPLEMENTAL MATERIAL

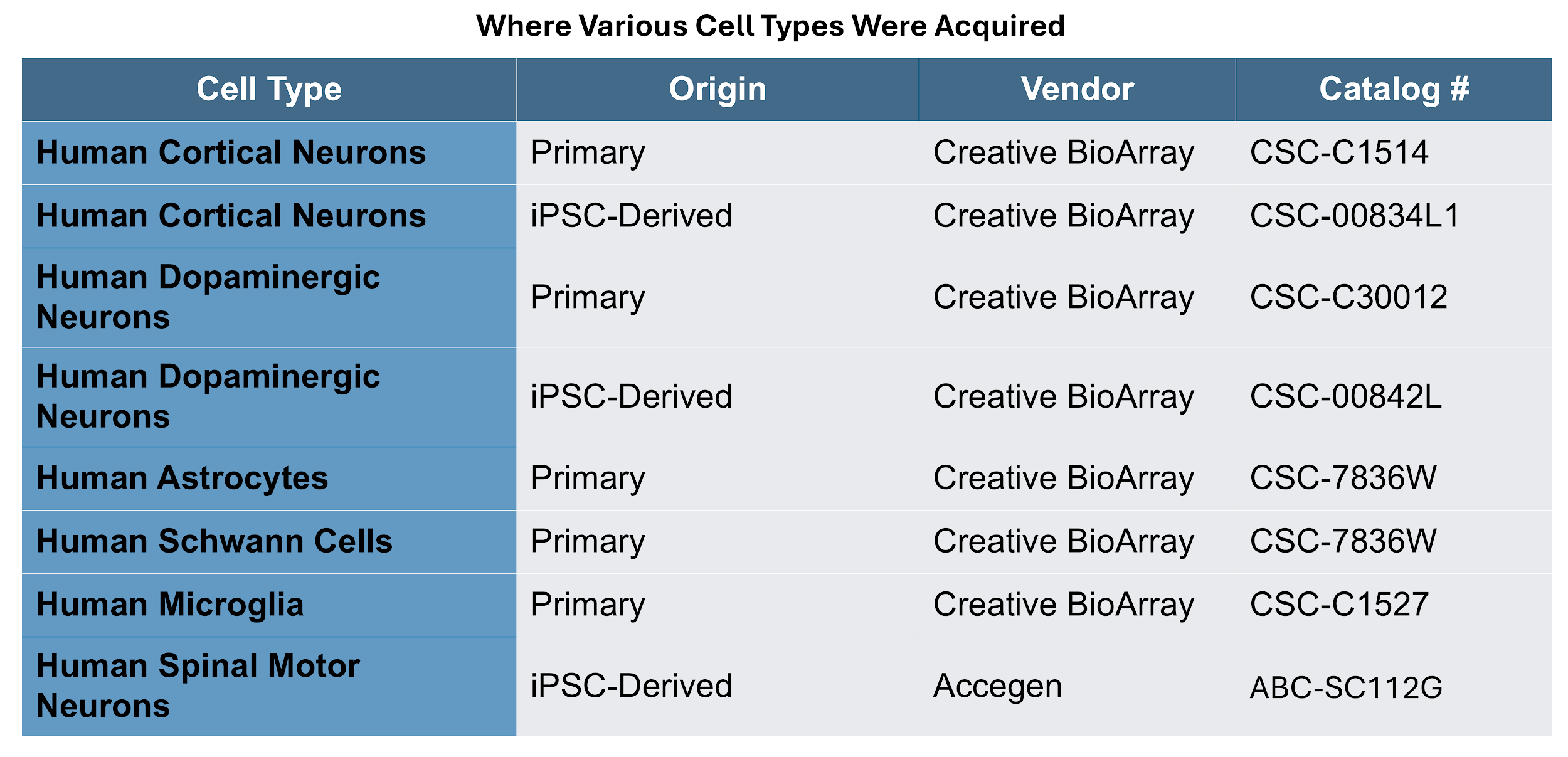

Supplemental Table 1 - Cell types used for model training and analytical validation, including cell origin (primary or iPSC-derived), vendor, and catalog numbers.

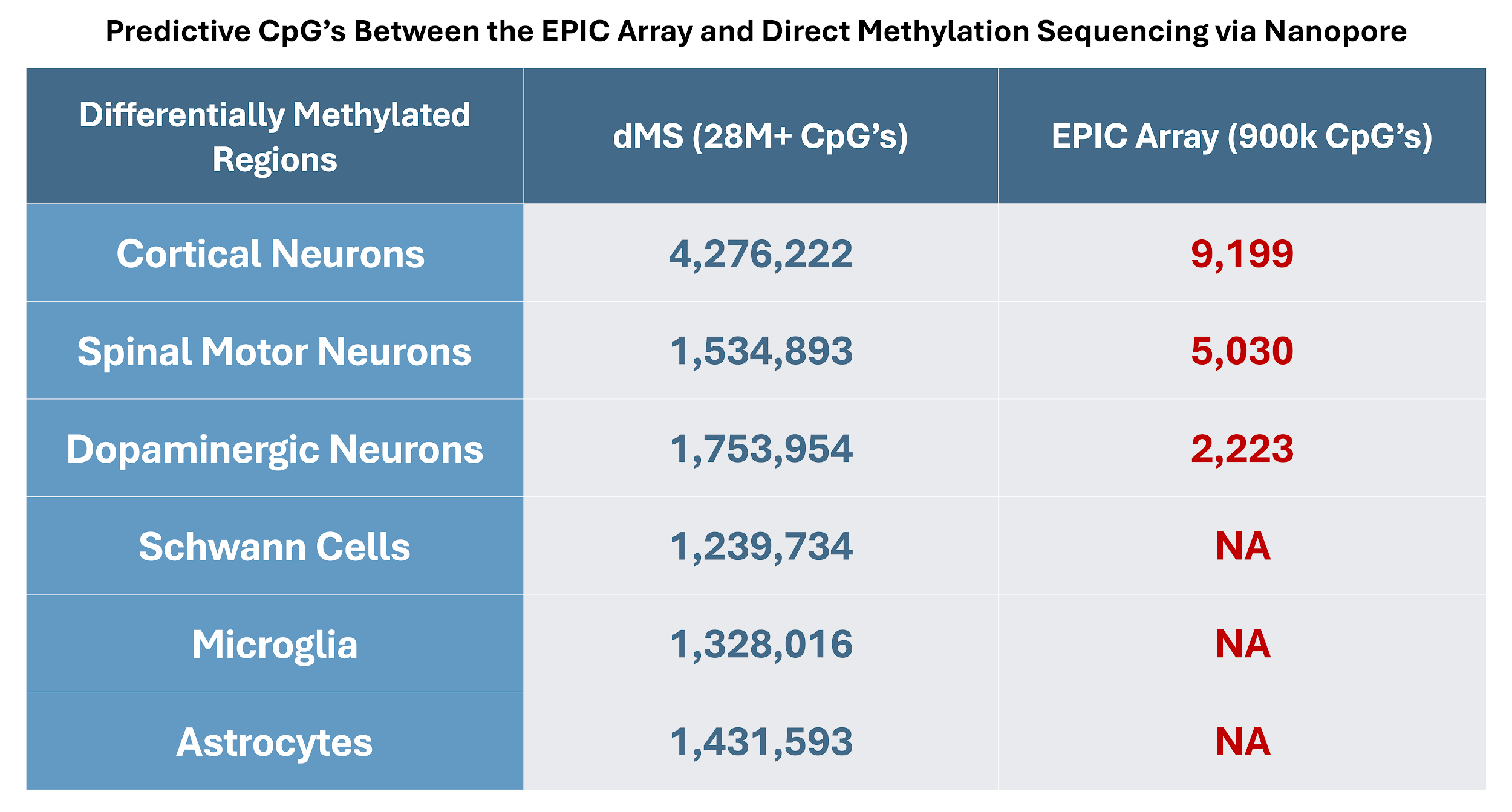

Supplemental Table 2 - Table summarizing the total number of unique DMRs identified per cell type using ONT-based native methylation sequencing, including a comparison to the coverage achievable using the Illumina EPIC array.

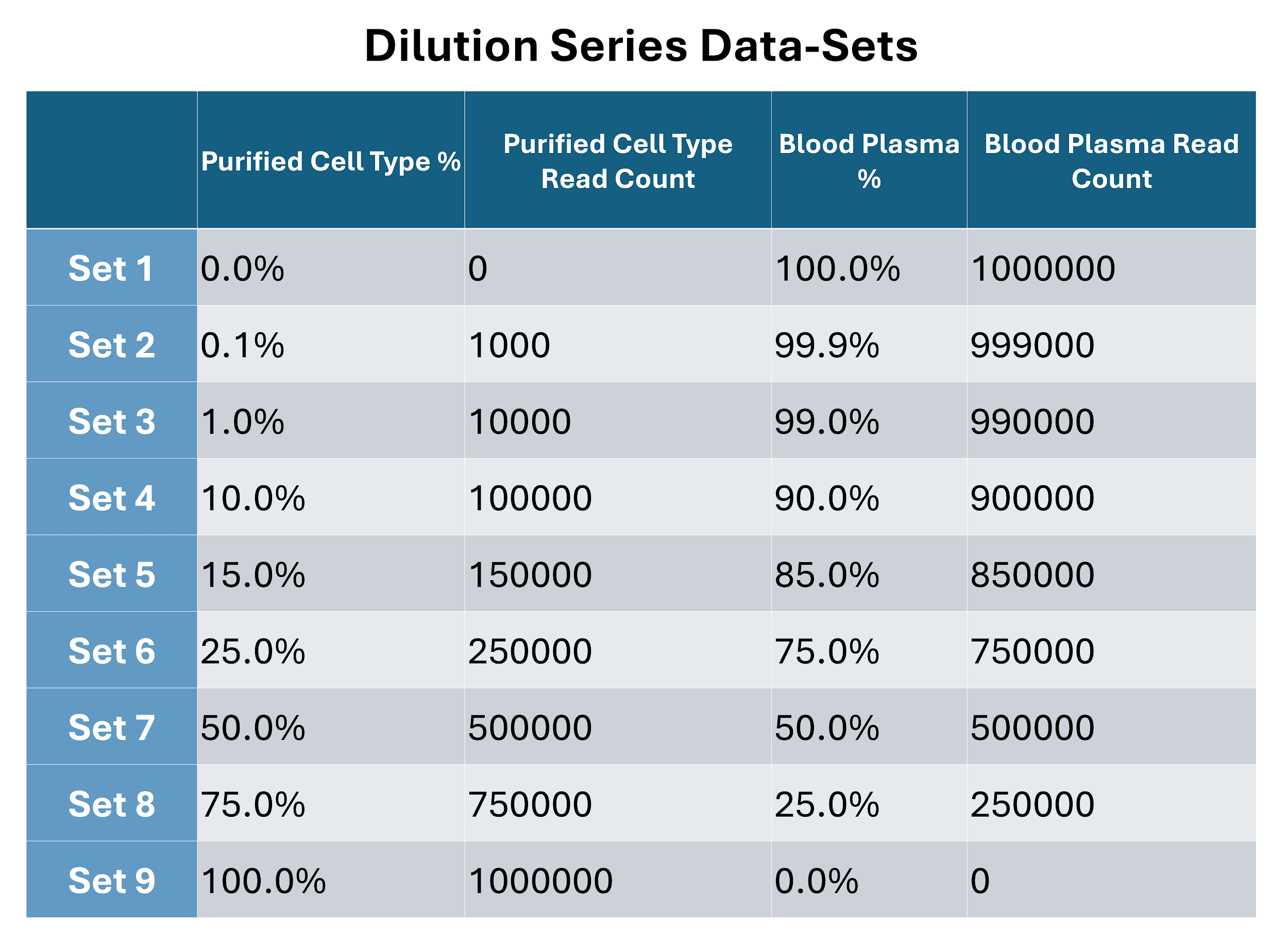

Supplemental Table 3 - Detailed breakdown of synthetic dilution datasets including the proportion and absolute number of cell-derived versus plasma-derived reads in each mixture. Used for analytical validation of classifier performance.

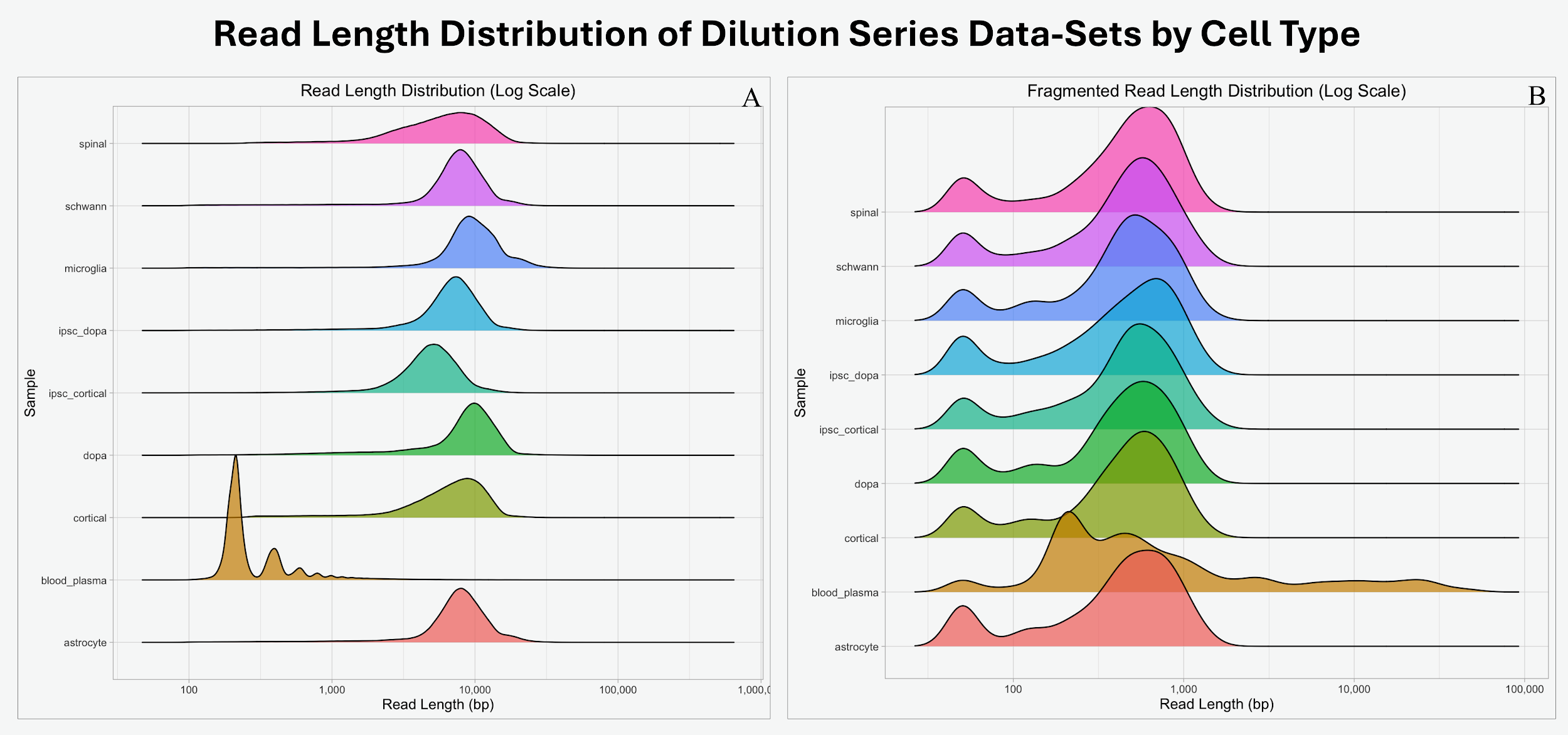

Supplemental Figure 1 - Histograms showing read length distributions for cell type-derived reads before and after synthetic fragmentation. (A) Read length distribution prior to synthetic fragmentation. (B) Read length distribution after synthetic fragmentation. This quality control step ensures that read lengths resemble true cfDNA distributions seen in plasma.

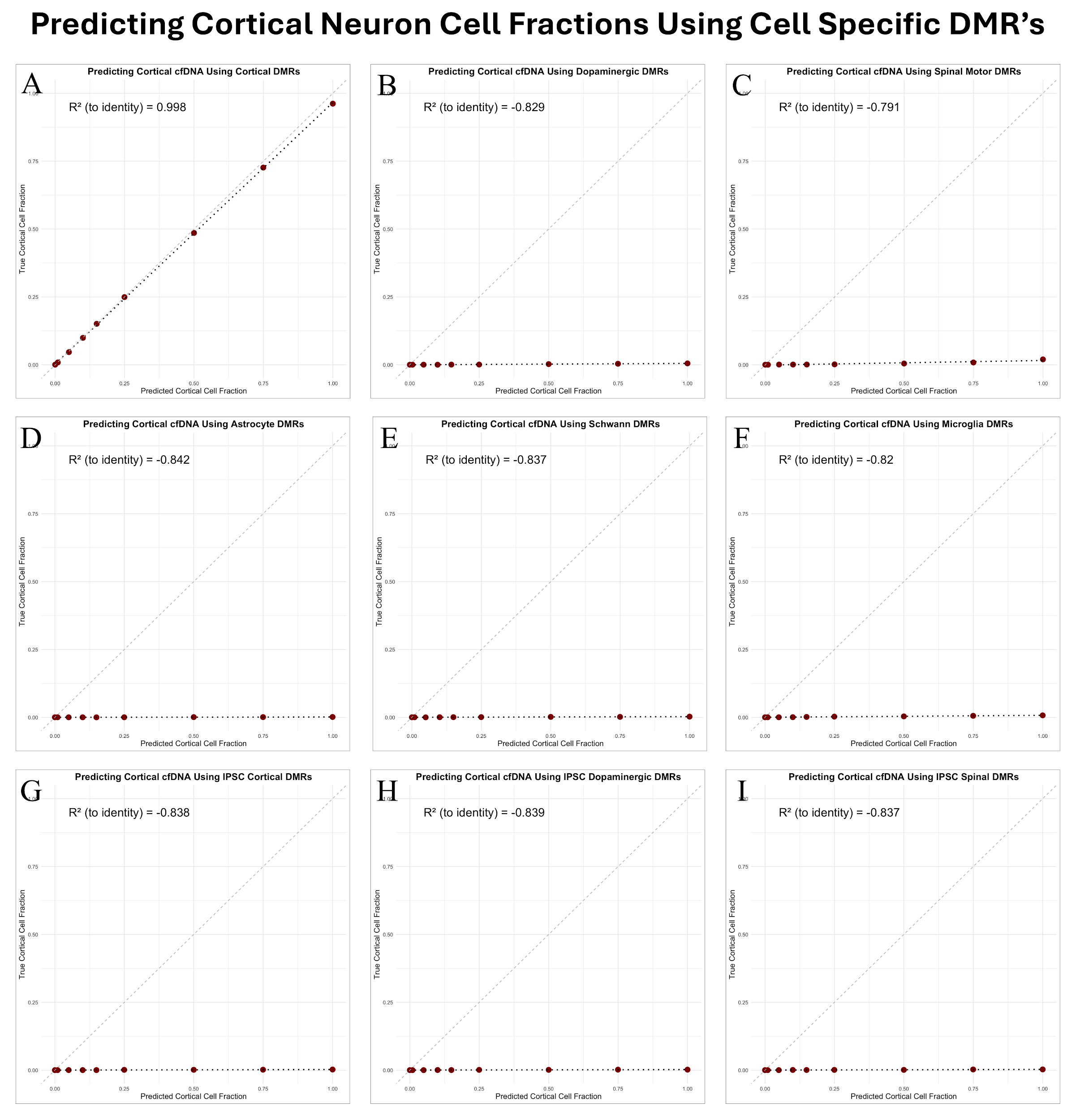

Supplemental Figure 2 - Correlation plots comparing actual vs predicted cortical neuron cell fractions in synthetic dilution datasets using various sets of established cell-specific DMRs. (A) Predicting the proportion of cortical derived cfDNA using cortical specific DMRs. (B) Predicting the proportion of cortical neuron derived cfDNA using dopaminergic neuron specific DMRs. (C) Predicting the proportion of cortical derived cfDNA using dopaminergic neuron specific DMRs. (D) Predicting the proportion of cortical derived cfDNA using astrocyte specific DMRs. (E) Predicting the proportion of cortical derived cfDNA using schwann cell specific DMRs. (F) Predicting the proportion of cortical derived cfDNA using microglial specific DMRs. (G) Predicting the proportion of cortical derived cfDNA using ipsc cortical neuron specific DMRs. (H) Predicting the proportion of cortical derived cfDNA using ipsc dopaminergic neuron specific DMRs. (I) Predicting the proportion of cortical derived cfDNA using ipsc spinal motor neuron specific DMRs.

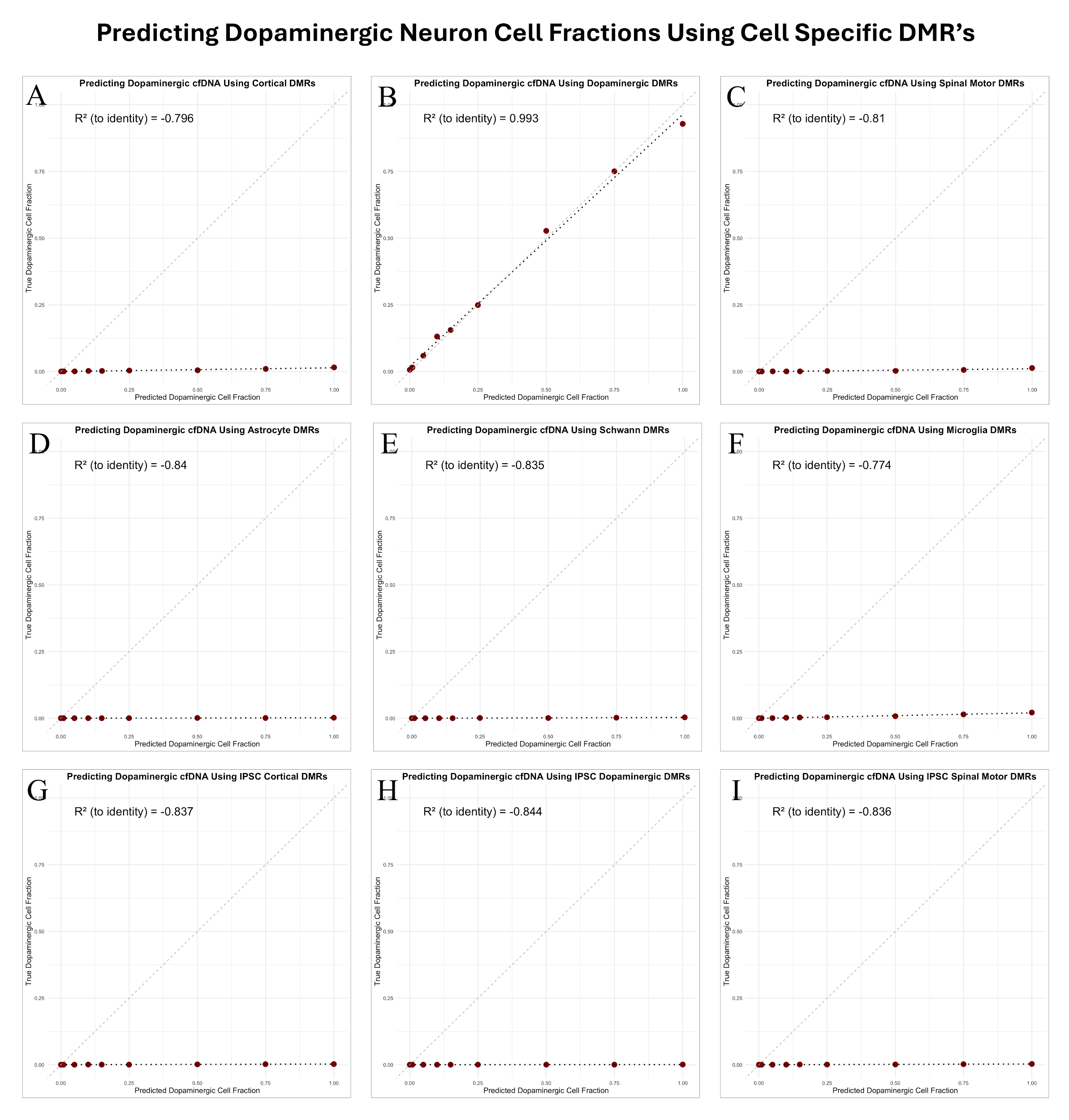

Supplemental Figure 3 - Correlation plots comparing actual vs predicted dopaminergic neuron cell fractions in synthetic dilution datasets using various sets of established cell-specific DMRs. (A) Predicting the proportion of dopaminergic derived cfDNA using cortical specific DMRs. (B) Predicting the proportion of dopaminergic neuron derived cfDNA using dopaminergic neuron specific DMRs. (C) Predicting the proportion of dopaminergic derived cfDNA using spinal motor neuron specific DMRs. (D) Predicting the proportion of dopaminergic derived cfDNA using astrocyte specific DMRs. (E) Predicting the proportion of dopaminergic derived cfDNA using schwann cell specific DMRs. (F) Predicting the proportion of dopaminergic derived cfDNA using microglial specific DMRs. (G) Predicting the proportion of dopaminergic derived cfDNA using ipsc cortical neuron specific DMRs. (H) Predicting the proportion of dopaminergic derived cfDNA using ipsc dopaminergic neuron specific DMRs. (I) Predicting the proportion of dopaminergic derived cfDNA using ipsc spinal motor neuron specific DMRs.

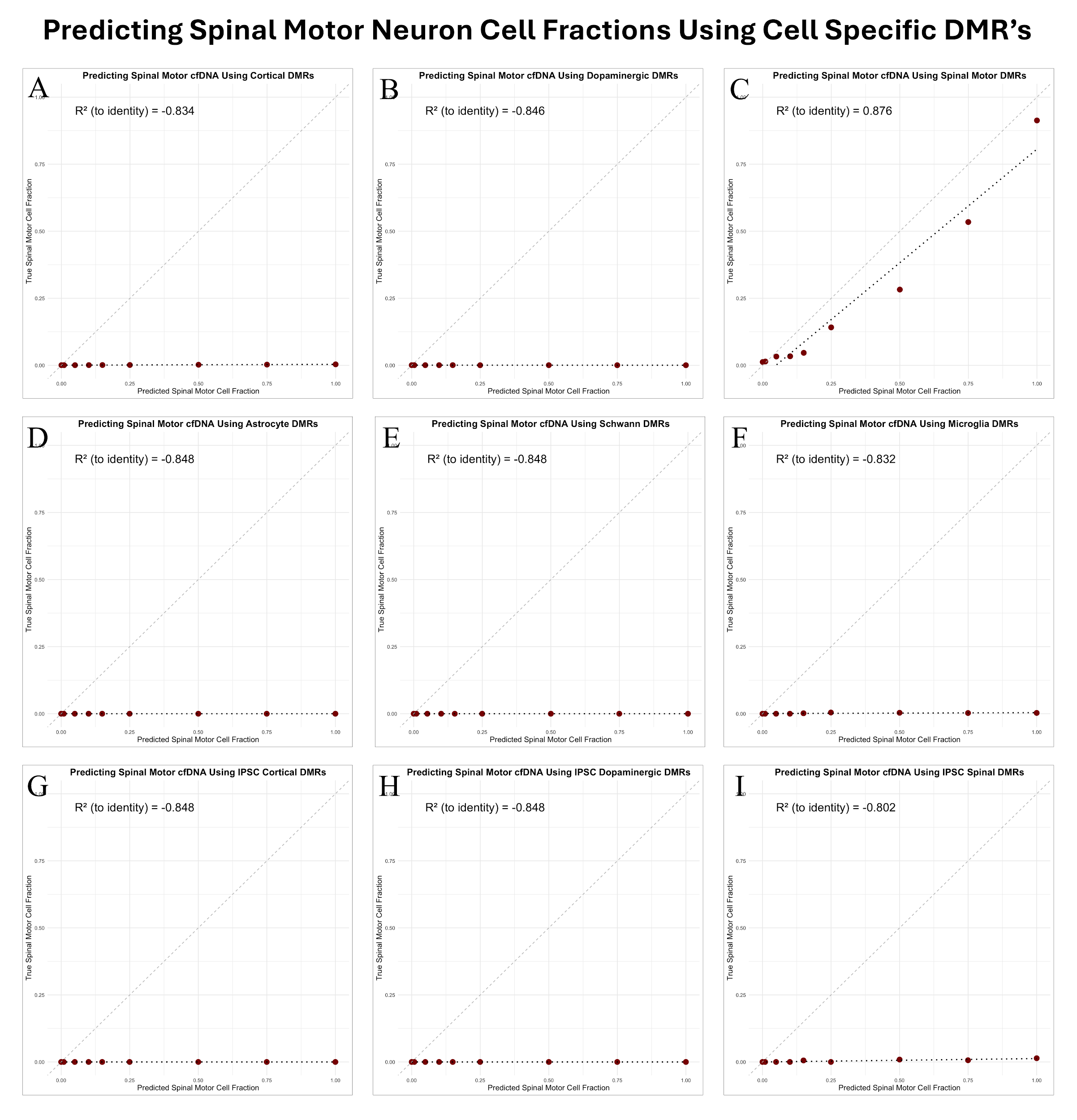

Supplemental Figure 4 - Correlation plots comparing actual vs predicted spinal motor neuron cell fractions in synthetic dilution datasets using various sets of established cell-specific DMRs. (A) Predicting the proportion of spinal motor derived cfDNA using cortical specific DMRs. (B) Predicting the proportion of spinal motor neuron derived cfDNA using dopaminergic neuron specific DMRs. (C) Predicting the proportion of spinal motor derived cfDNA using spinal motor neuron specific DMRs. (D) Predicting the proportion of spinal motor derived cfDNA using astrocyte specific DMRs. (E) Predicting the proportion of spinal motor derived cfDNA using schwann cell specific DMRs. (F) Predicting the proportion of spinal motor derived cfDNA using microglial specific DMRs. (G) Predicting the proportion of spinal motor derived cfDNA using ipsc cortical neuron specific DMRs. (H) Predicting the proportion of spinal motor derived cfDNA using ipsc dopaminergic neuron specific DMRs. (I) Predicting the proportion of spinal motor derived cfDNA using ipsc spinal motor neuron specific DMRs.

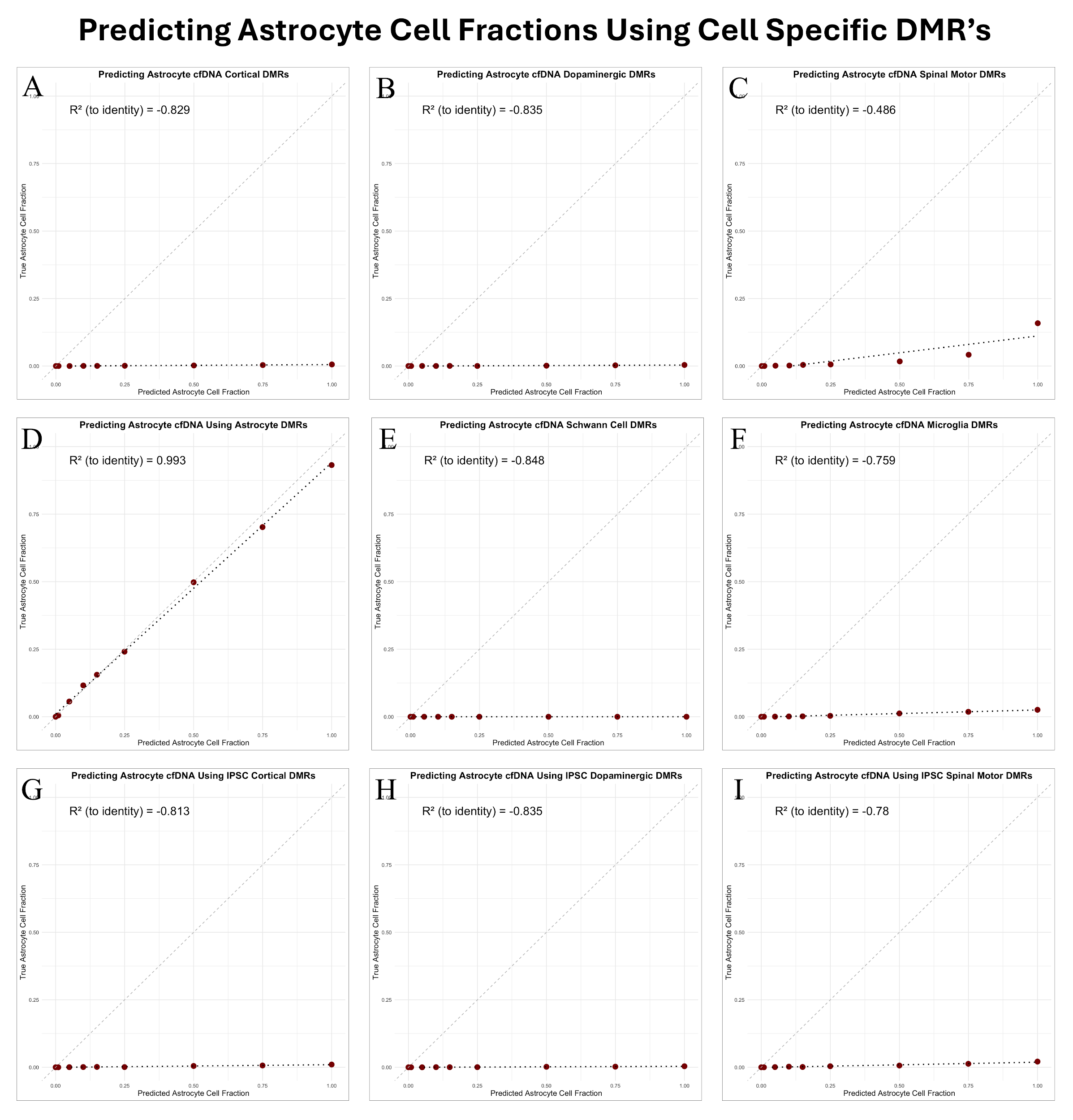

Supplemental Figure 5 - Correlation plots comparing actual vs predicted astrocyte cell fractions in synthetic dilution datasets using various sets of established cell-specific DMRs. (A) Predicting the proportion of astrocyte derived cfDNA using cortical specific DMRs. (B) Predicting the proportion of astrocyte neuron derived cfDNA using dopaminergic neuron specific DMRs. (C) Predicting the proportion of astrocyte derived cfDNA using spinal motor neuron specific DMRs. (D) Predicting the proportion of astrocyte derived cfDNA using astrocyte specific DMRs. (E) Predicting the proportion of astrocyte derived cfDNA using schwann cell specific DMRs. (F) Predicting the proportion of astrocyte derived cfDNA using microglial specific DMRs. (G) Predicting the proportion of astrocyte derived cfDNA using ipsc cortical neuron specific DMRs. (H) Predicting the proportion of astrocyte derived cfDNA using ipsc dopaminergic neuron specific DMRs. (I) Predicting the proportion of astrocyte derived cfDNA using ipsc spinal motor neuron specific DMRs.

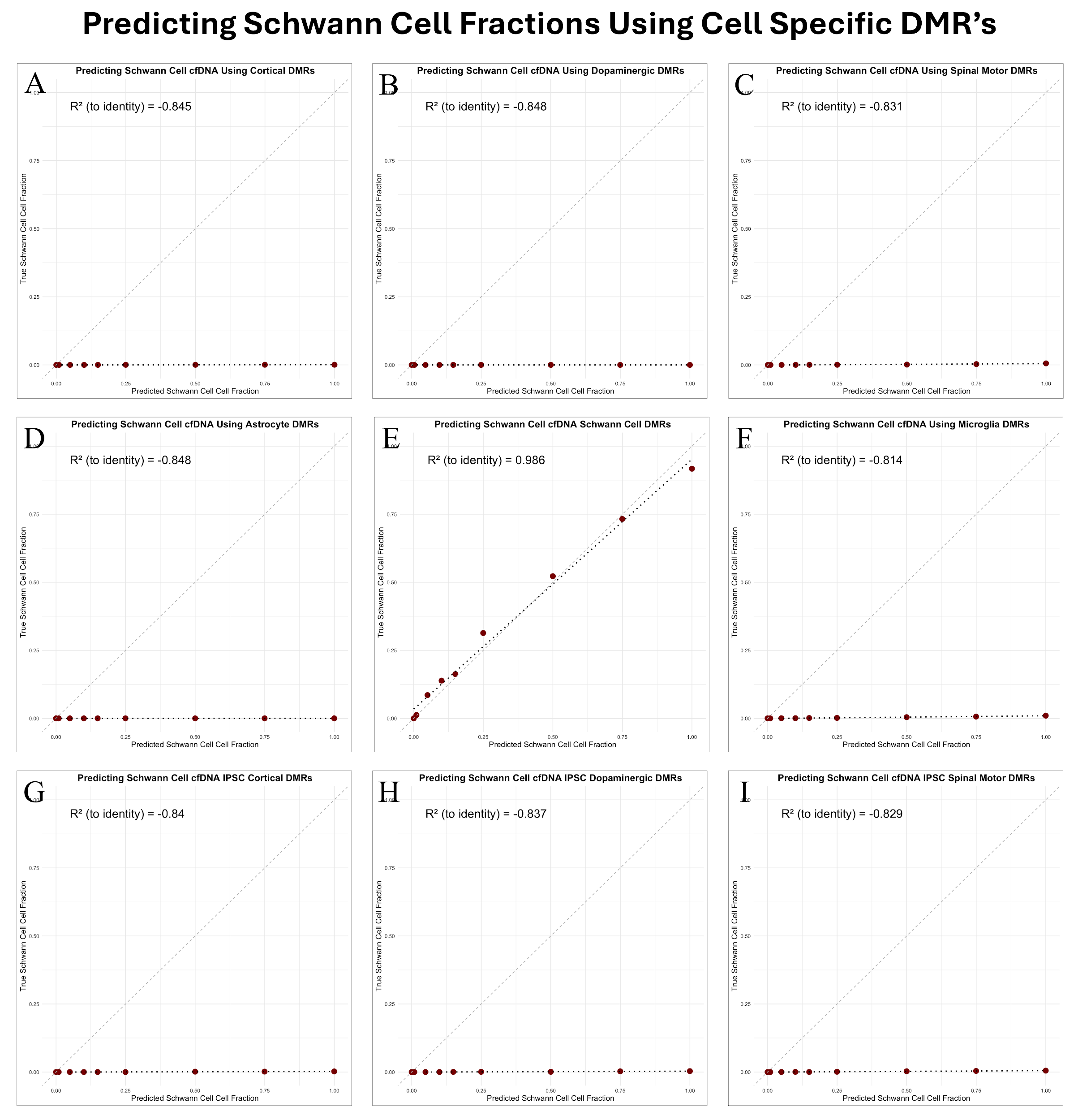

Supplemental Figure 6 - Correlation plots comparing actual vs predicted schwann cell fractions in synthetic dilution datasets using various sets of established cell-specific DMRs. (A) Predicting the proportion of schwann cell derived cfDNA using cortical specific DMRs. (B) Predicting the proportion of schwann cell neuron derived cfDNA using dopaminergic neuron specific DMRs. (C) Predicting the proportion of schwann cell derived cfDNA using spinal motor neuron specific DMRs. (D) Predicting the proportion of schwann cell derived cfDNA using astrocyte specific DMRs. (E) Predicting the proportion of schwann cell derived cfDNA using schwann cell specific DMRs. (F) Predicting the proportion of schwann cell derived cfDNA using microglial specific DMRs. (G) Predicting the proportion of schwann cell derived cfDNA using ipsc cortical neuron specific DMRs. (H) Predicting the proportion of schwann cell derived cfDNA using ipsc dopaminergic neuron specific DMRs. (I) Predicting the proportion of schwann cell derived cfDNA using ipsc spinal motor neuron specific DMRs.

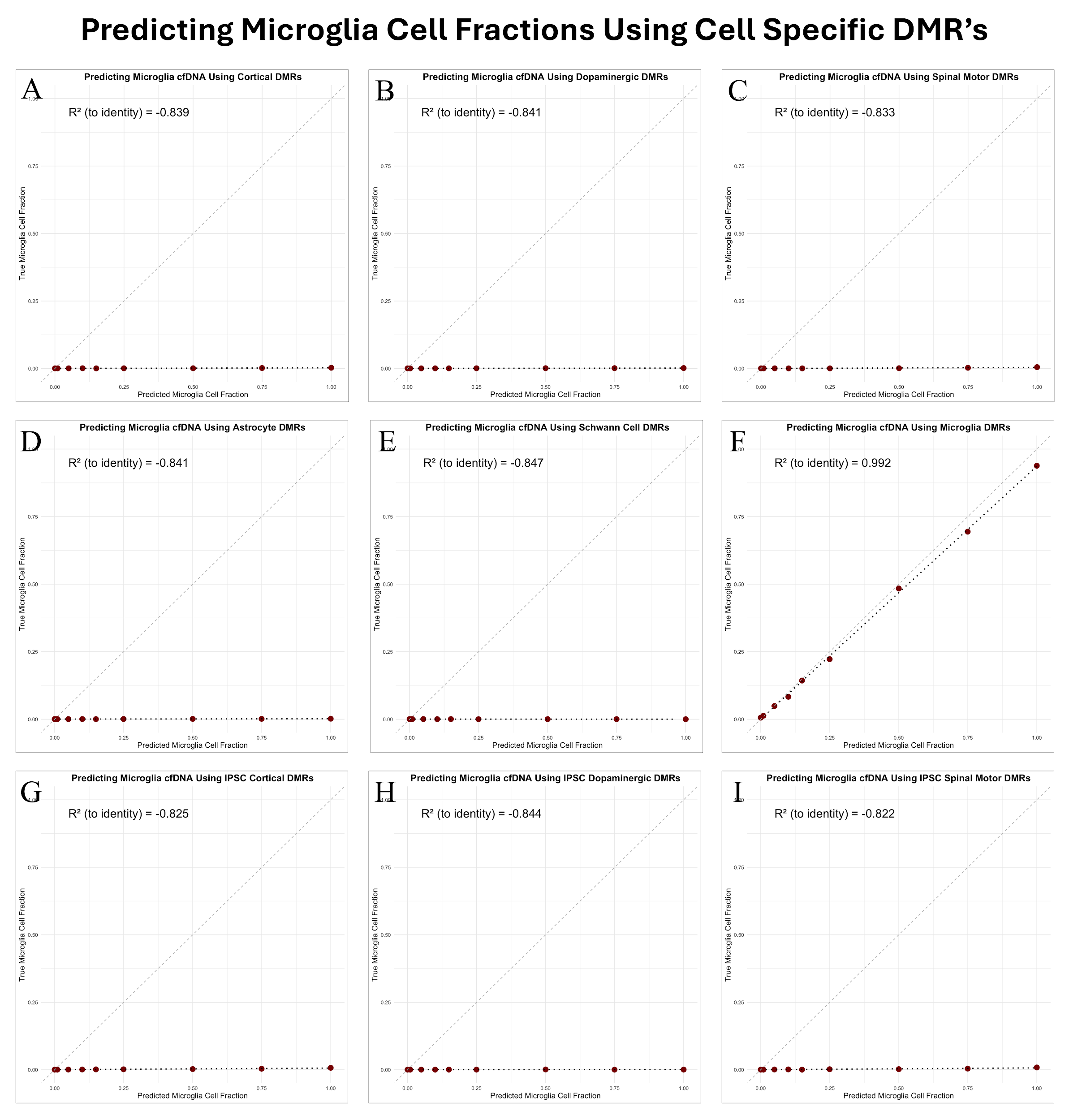

Supplemental Figure 7 - Correlation plots comparing actual vs predicted microglial fractions in synthetic dilution datasets using various sets of established cell-specific DMRs. (A) Predicting the proportion of microglia derived cfDNA using cortical specific DMRs. (B) Predicting the proportion of microglia neuron derived cfDNA using dopaminergic neuron specific DMRs. (C) Predicting the proportion of microglia derived cfDNA using spinal motor neuron specific DMRs. (D) Predicting the proportion of microglia derived cfDNA using astrocyte specific DMRs. (E) Predicting the proportion of microglia derived cfDNA using schwann cell specific DMRs. (F) Predicting the proportion of microglia derived cfDNA using microglial specific DMRs. (G) Predicting the proportion of microglia derived cfDNA using ipsc cortical neuron specific DMRs. (H) Predicting the proportion of microglia derived cfDNA using ipsc dopaminergic neuron specific DMRs. (I) Predicting the proportion of microglia derived cfDNA using ipsc spinal motor neuron specific DMRs.

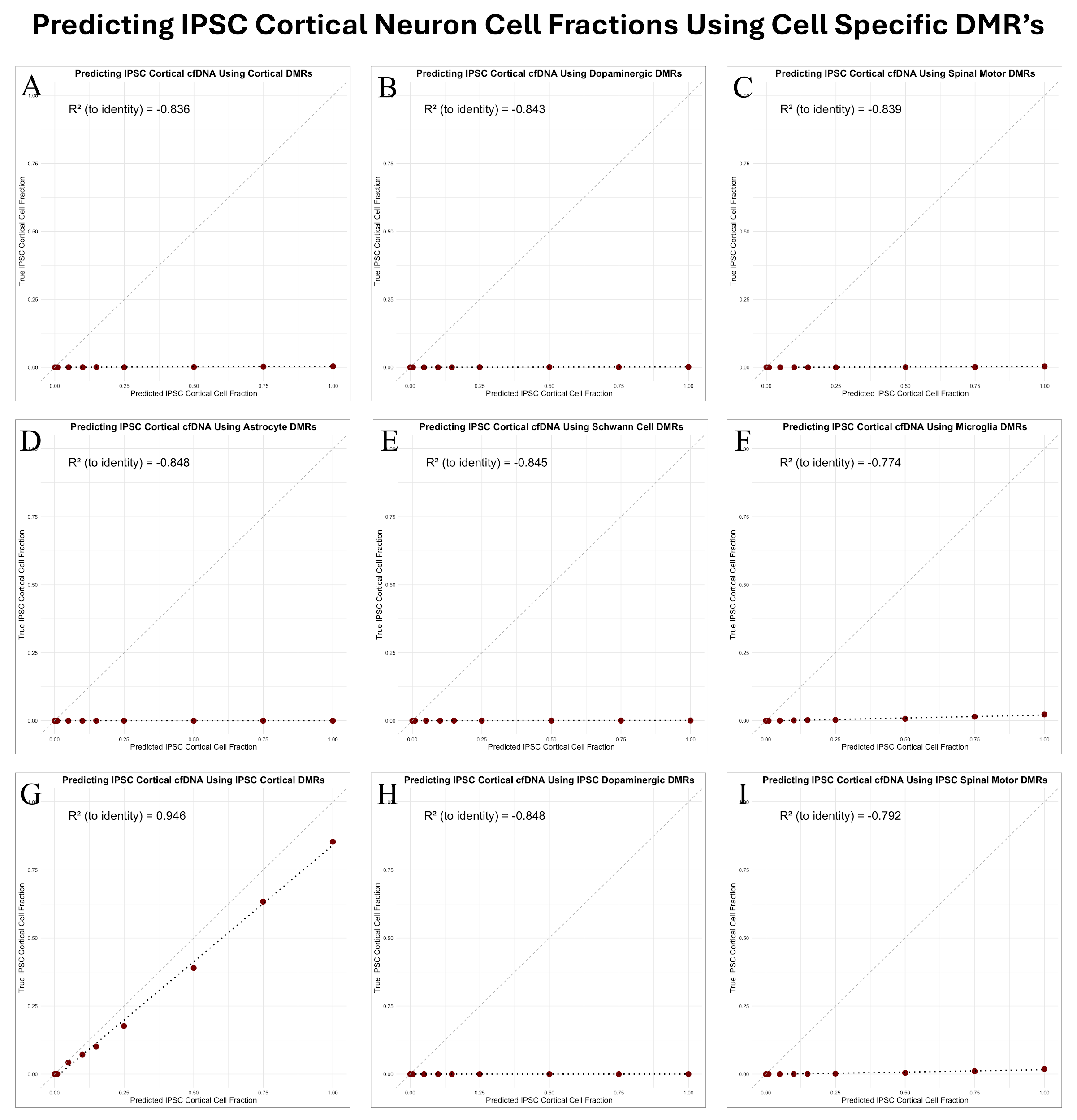

Supplemental Figure 8 - Correlation plots comparing actual vs predicted IPSC-derived cortical neuron fractions in synthetic dilution datasets using various sets of established cell-specific DMRs. (A) Predicting the proportion of IPSC cortical derived cfDNA using cortical specific DMRs. (B) Predicting the proportion of IPSC cortical neuron derived cfDNA using dopaminergic neuron specific DMRs. (C) Predicting the proportion of IPSC cortical derived cfDNA using spinal motor neuron specific DMRs. (D) Predicting the proportion of IPSC cortical derived cfDNA using astrocyte specific DMRs. (E) Predicting the proportion of IPSC cortical derived cfDNA using schwann cell specific DMRs. (F) Predicting the proportion of IPSC cortical derived cfDNA using microglial specific DMRs. (G) Predicting the proportion of IPSC cortical derived cfDNA using IPSC cortical neuron specific DMRs. (H) Predicting the proportion of IPSC cortical derived cfDNA using IPSC dopaminergic neuron specific DMRs. (I) Predicting the proportion of IPSC cortical derived cfDNA using IPSC spinal motor neuron specific DMRs.

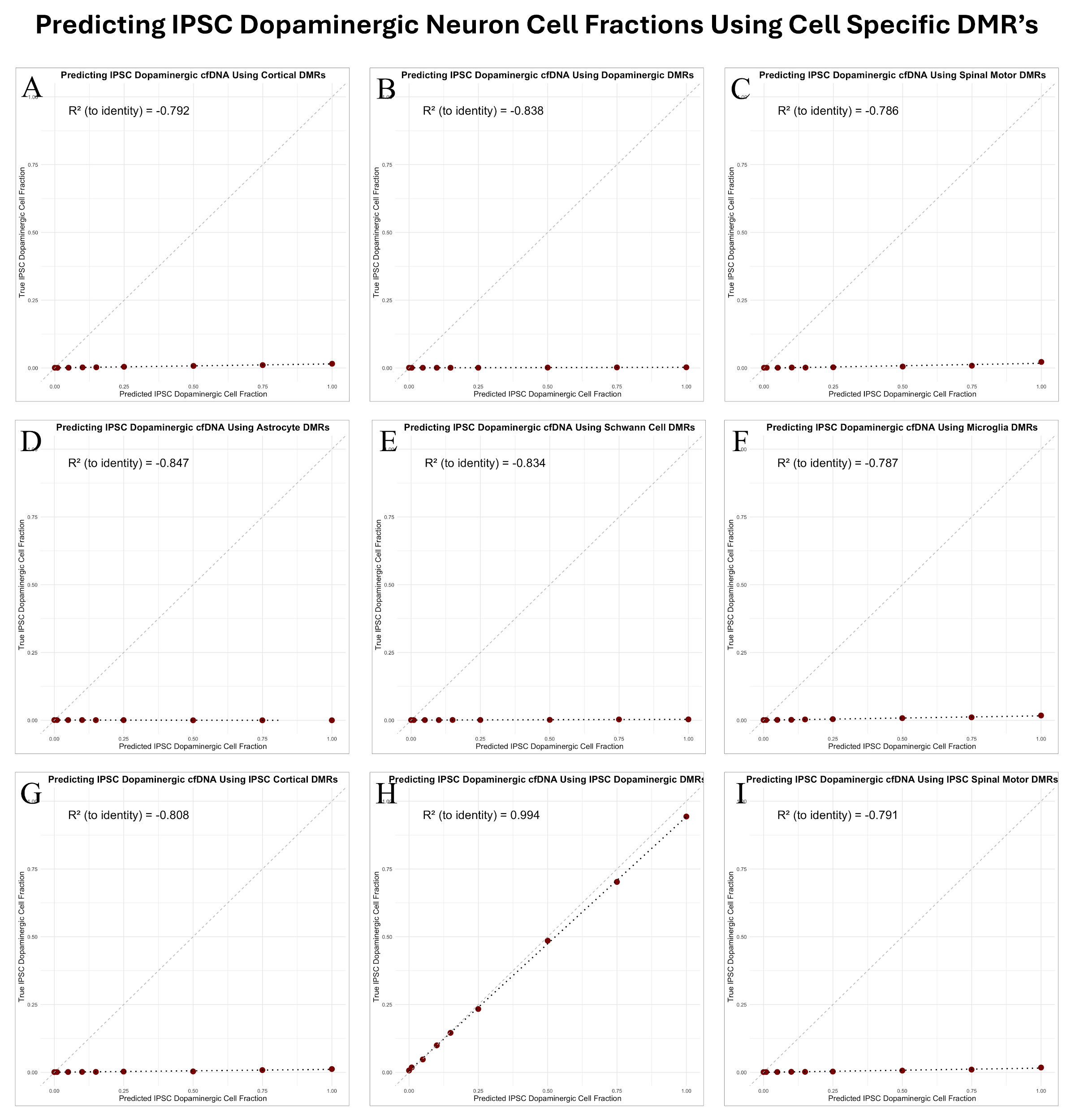

Supplemental Figure 9 - Correlation plots comparing actual vs predicted IPSC-derived dopaminergic neuron fractions in synthetic dilution datasets using various sets of established cell-specific DMRs. (A) Predicting the proportion of IPSC dopaminergic derived cfDNA using cortical specific DMRs. (B) Predicting the proportion of IPSC dopaminergic neuron derived cfDNA using dopaminergic neuron specific DMRs. (C) Predicting the proportion of IPSC dopaminergic derived cfDNA using spinal motor neuron specific DMRs. (D) Predicting the proportion of IPSC dopaminergic derived cfDNA using astrocyte specific DMRs. (E) Predicting the proportion of IPSC dopaminergic derived cfDNA using schwann cell specific DMRs. (F) Predicting the proportion of IPSC dopaminergic derived cfDNA using microglial specific DMRs. (G) Predicting the proportion of IPSC dopaminergic derived cfDNA using IPSC cortical neuron specific DMRs. (H) Predicting the proportion of IPSC dopaminergic derived cfDNA using IPSC dopaminergic neuron specific DMRs. (I) Predicting the proportion of IPSC dopaminergic derived cfDNA using IPSC spinal motor neuron specific DMRs.

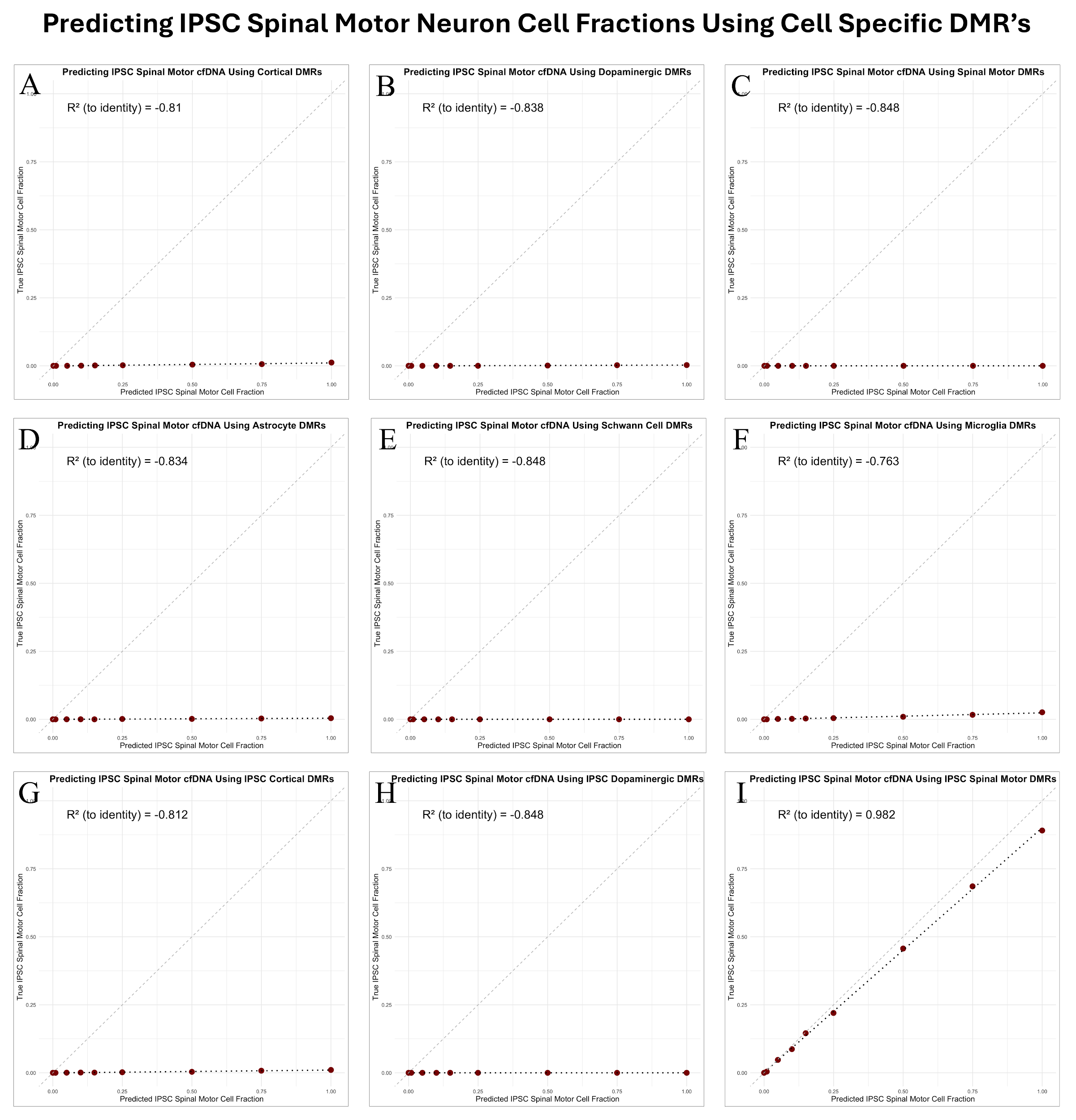

Supplemental Figure 10 - Correlation plots comparing actual vs predicted IPSC-derived spinal motor neuron fractions in synthetic dilution datasets using various sets of established cell-specific DMRs. (A) Predicting the proportion of IPSC spinal motor derived cfDNA using cortical specific DMRs. (B) Predicting the proportion of IPSC spinal motor neuron derived cfDNA using dopaminergic neuron specific DMRs. (C) Predicting the proportion of IPSC spinal motor derived cfDNA using spinal motor neuron specific DMRs. (D) Predicting the proportion of IPSC spinal motor derived cfDNA using astrocyte specific DMRs. (E) Predicting the proportion of IPSC spinal motor derived cfDNA using schwann cell specific DMRs. (F) Predicting the proportion of IPSC spinal motor derived cfDNA using microglial specific DMRs. (G) Predicting the proportion of IPSC spinal motor derived cfDNA using IPSC cortical neuron specific DMRs. (H) Predicting the proportion of IPSC spinal motor derived cfDNA using IPSC dopaminergic neuron specific DMRs. (I) Predicting the proportion of IPSC spinal motor derived cfDNA using IPSC spinal motor neuron specific DMRs.

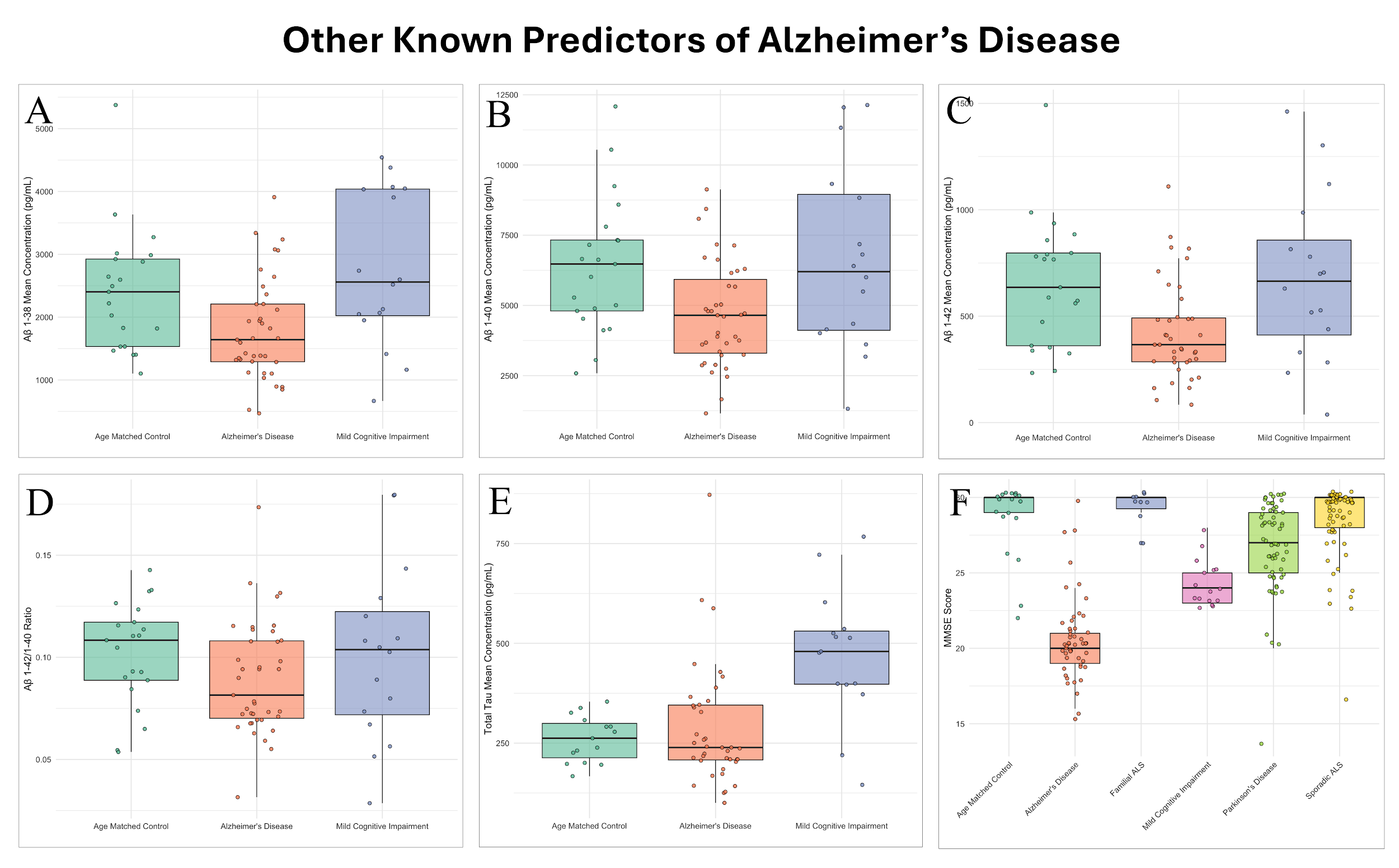

Supplemental Figure 11 - Box plots showing traditional biomarkers of Alzheimer’s disease in the study cohort. These include (A) amyloid-beta 1-38, (B)1-40, and (C) 1-42 concentrations, the (D) 1-42/1-40 ratio, (E) total tau, and (F) MMSE scores. This figure serves as a clinical comparator, highlighting the limited discriminative power of protein biomarkers relative to cfDNA methylation signals.

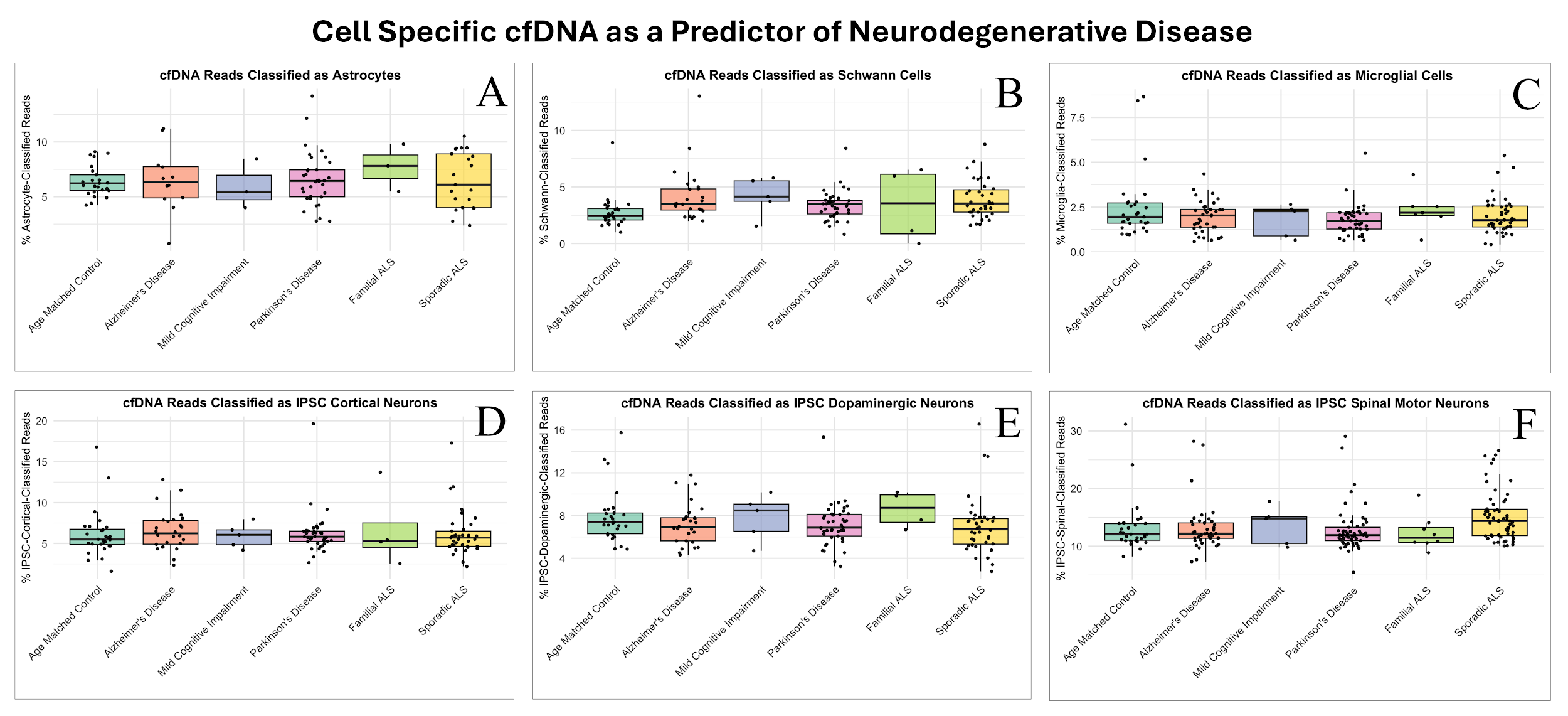

Supplemental Figure 12 - Box plots illustrate the percentage of classified cfDNA from non-cortical, non-dopaminergic, and non-spinal motor cell types across all clinical cohorts. Cell types include (A) astrocytes, (B) Schwann cells, (C) microglia, (D) iPSC cortical, (E) iPSC Dopaminergic, and (F) iPSC spinal motor neurons. These plots emphasize the specificity of signal to disease-relevant cell types and further validate the limited utility of iPSC-derived models.

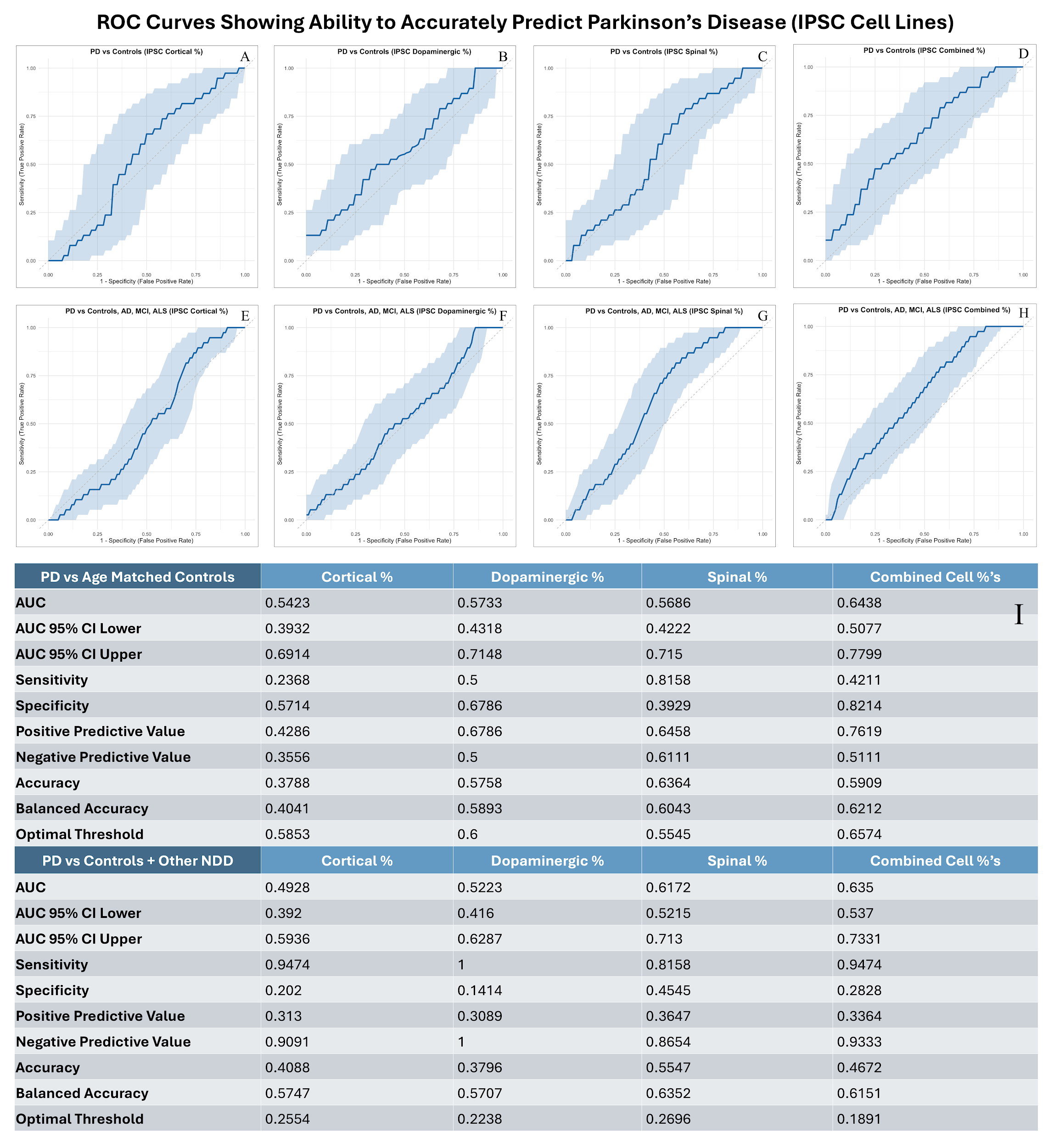

Supplemental Figure 13 - Receiver Operating Characteristic (ROC) curves for predicting Alzheimer’s Disease using IPSC derived models for (a)(e) cortical, (b)(f) dopaminergic, and (c)(g) spinal neuron-derived cfDNA, alone and in (d)(h) combination. Panels compare classification performance against both controls and other neurodegenerative diseases, showcasing the additive value of combining multiple cell type signals.

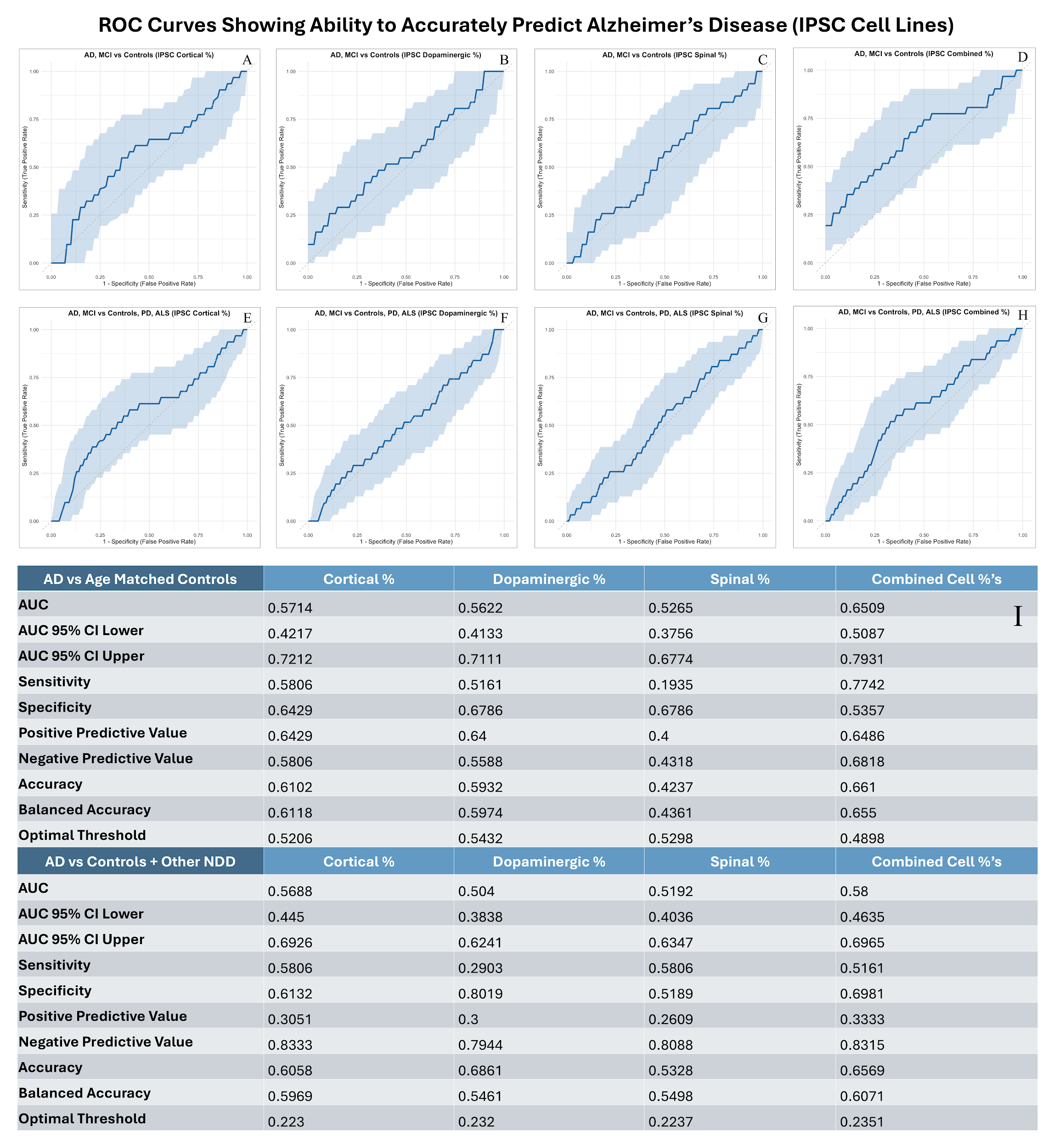

Supplemental Figure 14 - Receiver Operating Characteristic (ROC) curves for predicting Parkinson’s Disease using IPSC derived models for (a)(e) cortical, (b)(f) dopaminergic, and (c)(g) spinal neuron-derived cfDNA, alone and in (d)(h) combination. Panels compare classification performance against both controls and other neurodegenerative diseases, showcasing the additive value of combining multiple cell type signals.

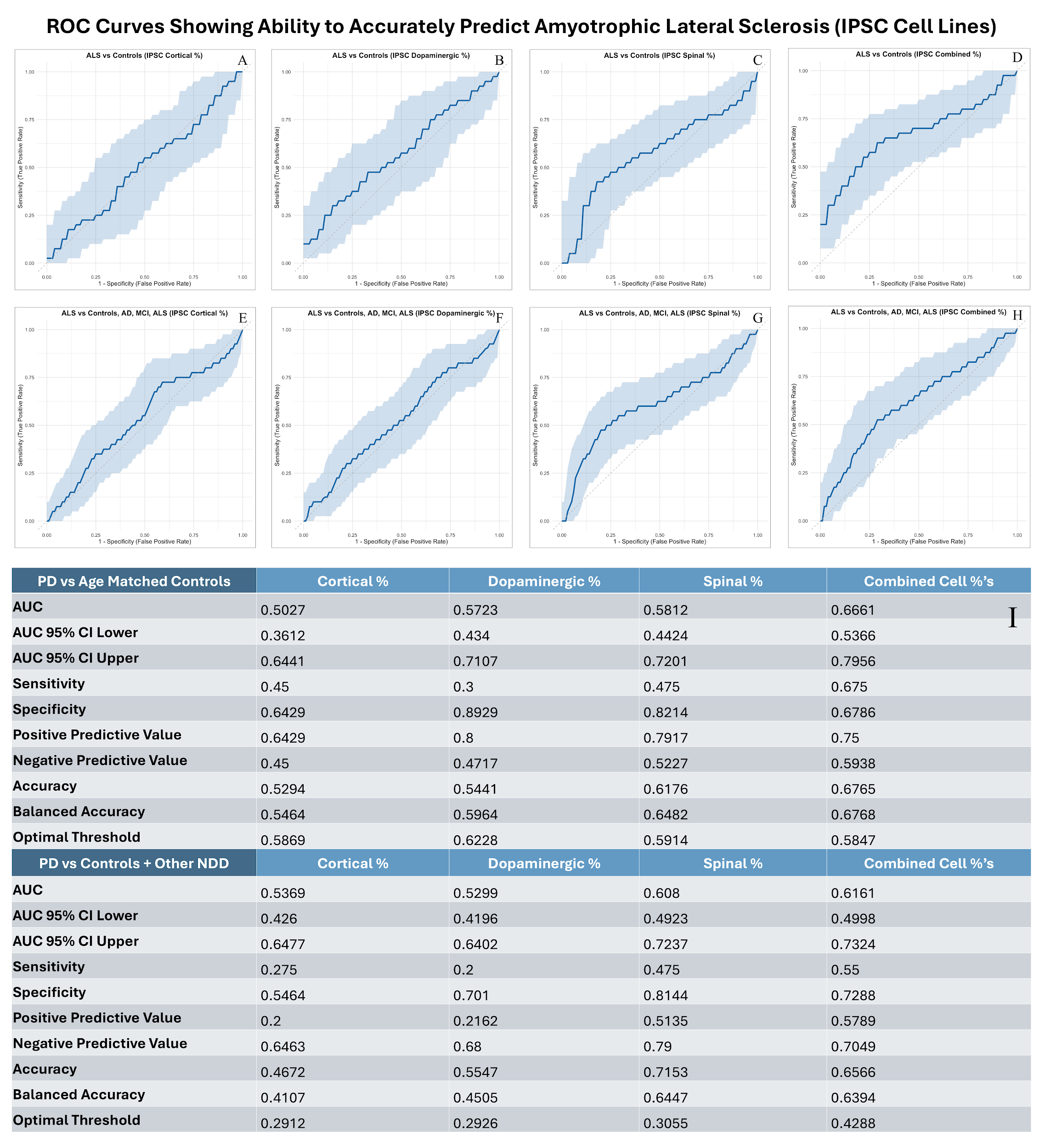

Supplemental Figure 15 - Receiver Operating Characteristic (ROC) curves for predicting Amyotrophic Lateral Sclerosis using IPSC derived models for (a)(e) cortical, (b)(f) dopaminergic, and (c)(g) spinal neuron-derived cfDNA, alone and in (d)(h) combination. Panels compare classification performance against both controls and other neurodegenerative diseases, showcasing the additive value of combining multiple cell type signals.
